# Patterns of HIV-1 viral load suppression and drug resistance during the dolutegravir transition: a population-based longitudinal study

**DOI:** 10.1101/2025.09.01.25334862

**Authors:** Michael A. Martin, Alexandra Blenkinsop, Michelle Moffa, Steven James Reynolds, Fred Nalugoda, Thomas C. Quinn, Godfrey Kigozi, Robert Ssekubugu, Ravindra K. Gupta, Nicholas E. Grayson, George MacIntyre-Cockett, Joseph Kagaayi, Gertrude Nakigozi, Lucie Abeler-Dörner, Christophe Fraser, Oliver Ratmann, Aaron A.R. Tobian, Oliver Laeyendecker, Sikhulile Moyo, Caitlin E. Kennedy, David Bonsall, Ronald Moses Galiwango, M. Kate Grabowski, the Rakai Health Sciences Program, the PANGEA-HIV Consortium

## Abstract

**Background:** Data on the population-scale impact of dolutegravir (DTG)-based HIV regimens in sub-Saharan Africa are extremely limited. We used data from a surveillance cohort in southern Uganda to assess viral suppression and antiretroviral (ART) resistance over 10-years alongside DTG scale-up.

**Methods:** Consenting participants in the population-based Rakai Community Cohort Study between August 2011 and March 2023 aged 15-59 completed questionnaires and provided samples for HIV testing, viral load quantification, and viral deep-sequencing. We collected data on DTG-utilization at HIV care clinics. We estimated the prevalence of HIV suppression (<1,000 copies/mL) and ART resistance using robust Poisson regression. Bayesian logistic regression quantified associations between resistance and individual-level suppression across surveys.

**Findings:** Among 20,383 people living with HIV (PLHIV), suppression increased from 57.1% (95% confidence interval [CI]: 55.4%-58.8%) to 90.3% (95%CI: 89.2%-91.4%) between 2014 and 2022. By 2020 84.4% (95%CI: 83.7%-85.2%) and 64.6% (95%CI: 63.9%-65.3%) of men and women were on DTG regimens. Among treatment-experienced viremic PLHIV, overall resistance decreased from 51.1% (95%CI: 40.7%-64.1%, 2014) to 27.9% (95%CI: 21.3%-36.5%, 2022). Only two participants harbored intermediate/high-level DTG resistance, attributable to inQ148R, inE138K, and inG140A. Low-level INSTI resistance (inS153Y) was observed in 23/207 (7.5%) of viremic individuals, with putative evidence of transmission. By 2022, suppression was unrelated to prior history of NNRTI/NRTI resistance (risk ratios: 1.14, 95%HPD: 0.96-1.32 and 1.12, 95%HPD: 0.88 - 1.35).

**Interpretation:** Viral suppression increased during the DTG-transition with minimal emerging intermediate/high-level resistance. Falling resistance among treatment-experienced PLHIV underscores the role of ART adherence in reducing viremia. The emergence of inS153Y justifies continued genomic surveillance of ART resistance.

**Funding:** National Institutes of Health and the Gates Foundation.

**Research in context:** *Evidence before the study:* We searched PubMed for studies matching the keywords “HIV” “resistance” “cohort” “dolutegravir” published after 2018, when dolutegravir (DTG) was first recommended for first-line use globally, and identified 108 studies. We excluded 78 studies, one for being a pure modeling study, one for being about HIV-2, two for being duplicates, two for being study protocols, five for evaluating DTG efficacy as a second, not first, line regimen, 11 for not including any data on individuals on DTG, 17 for focusing on a single sub-population (e.g. children or seniors), 18 for evaluating DTG two (as opposed to three)-drug regimens, and 21 for not having relevant outcomes (e.g. insulin sensitivity). While not indexed on PubMed, we analyzed the World Health Organization HIV Drug Resistance Brief Report 2024 along with the 30 studies from our targeted search. Among the remaining 30 studies, 27 were primary research articles and the remainder reviews in addition to the WHO report. Among the primary research articles, DTG-based first-line regimens were shown to be associated with high-levels of viral suppression among both ART initiators (e.g. 83.0% in South Africa and 84.6% in Tanzania) and those transitioning to DTG from other regimens (e.g. 90.5% in South Africa, 93.8% in Uganda, and 98.4% in Lesotho). Among the four countries in the Africa region reporting to the WHO, all reported levels of viral load suppression among adults receiving ART of >90% between 2019 and 2022, however, levels were not systematically higher among those on DTG, as opposed to NNRTI-based regimens. Among two studies reporting on pre-treatment non-nucleoside reverse transcriptase inhibitor (NNRTI) resistance in the DTG-era, it continued to increase and reached 14.3% (*n*=14) and 15.3% (*n*=137) in Tanzania and Zimbabwe, respectively. Pre-existing nucleoside reverse transcriptase inhibitor (NRTI) resistance, particularly rtM184V, was associated with DTG failure in 3/4 studies in which it was reported. Among 14 studies evaluating persons failing DTG therapy, emergent DTG resistance was generally rare, on the order of 0-10%, depending on setting. Among treatment-experienced individuals who failed DTG treatment in Mozambique, however, DTG resistance was more common (19.6%, 36/183). Further, another study based in South Africa reported that 60.3% (41/68) of people failing DTG therapy harbored intermediate/high-level resistance. Across studies, the most commonly reported emergent DTG-resistance conferring mutations were, E138K, G190A, Q148H/K/R, N155H/D, and R263K.

*Added value of this study:* Among existing real-world studies, all were clinic-based in design, meaning they enrolled PLHIV reporting to care clinics. As not all PLHIV are engaged or retained in care, the results of these studies may not be generalizable to the broader population of PLHIV. Further, in the absence of accurate denominators on the total number of people living with and without HIV, clinic-based studies are unable to accurately assess the real-world population-scale impact of interventions such as changes to first-line treatment regimens. Further, the reviewed studies focused solely on data collected during the transition to DTG-based regimens and are therefore unable to evaluate changes in population-scale virological outcomes during DTG scale-up in light of ongoing trends towards increasing rates of treatment initiation and suppression due to scale-up of global treatment and prevention programs. In the current study, we address these limitations by evaluating population-scale real-world virological outcomes during DTG scale-up in southern Uganda using data from the population-based Rakai Community Cohort Study. We found that the population-prevalence of viral load suppression among PLHIV increased from 86.1% to 89.4% concurrent with the DTG transition. We further observe a trend towards lower rates of NNRTI and NRTI resistance among those who remain viremic despite self-reporting being on treatment alongside increased rates of suppression among those with resistance. This suggests a shift in the population of viremic treatment-experienced PLHIV away from those who remain viremic because of resistance and towards those who are disengaged from care, which is not apparent from sampling only care-seeking PLHIV. Only two viremic individuals harbored intermediate/high-level DTG resistance. We also show a continued increase in pre-treatment NNRTI resistance despite discontinuation of NNRTI-based regimens, reaching 14.8% by 2022. Encouragingly, no pretreatment intermediate/high-level DTG resistance was observed and only two people with treatment experience harbored such resistance. However, a low-level INSTI resistance mutation, inS153Y, was identified in 7.5% (23/307) of sequenced PLHIV and genetic clustering indicates potential transmission of this mutation among 5 of these individuals.

*Implications of the available evidence:* The transition to DTG-based first-line regimens has supported continued increases in the population prevalence of HIV viral load suppression with limited evidence of emergent intermediate or high-level drug resistance thus far. Given minimal resistance, initiating pretreatment PLHIV on therapy and engaging disengaged treatment-experienced PLHIV are critical for continued progress towards HIV treatment milestones. Continued surveillance for resistance mutations is needed in light of increasing rates of resistance to NNRTIs, which are used in long-lasting injectable ART regimens, and for emerging novel INSTI resistance mutations.

## Introduction

Antiretroviral therapy (ART) suppresses Human immunodeficiency Virus (HIV) viral load, slowing disease progression^1^ and preventing viral transmission.^2^ Due to HIV’s high mutation rate, ART resistance conferring mutations can arise in people living with HIV (PLHIV) during treatment. These mutations restore viral fitness while on ART, reducing treatment efficacy. Combination ART increases the barrier to resistance as mutations generally don’t confer resistance to all components and the likelihood of multiple mutations arising within-individual is low.^3^ Nevertheless, resistance can arise, particularly during intermittent ART adherence.^4,5^ While PLHIV remain viremic, resistance mutations can be transmitted to seronegative contacts (transmitted resistance),^6^ increasing treatment failure rates.^7^

Prior to 2018 the World Health Organization (WHO) recommended efavirenz (EFV, a non-nucleoside reverse transcriptase inhibitor, NNRTI) given in combination with two nucleoside reverse transcriptase inhibitors (NRTIs) as first-line HIV treatment regimens.^8^ During the scale-up of HIV treatment and prevention programs there has been a marked increase in the worldwide prevalence of pre-treatment NNRTI resistance,^9–11^ due to the low genetic barrier to resistance,^12^ and by 2018 at least 18 WHO monitored countries had crossed the 10% threshold for triggering changes to first-line regimens.^13^

In 2018, the WHO recommended dolutegravir (DTG)-based regimens, an integrase strand transfer inhibitor (INSTI), as first-line HIV treatment.^14^ DTG is well-tolerated with minimal side effects,^15,16^ more effective than NNRTI/NRTI at achieving suppression in trials,^15,17,18^ and is thought to have a higher genetic barrier to resistance.^19^ By July 2024, 118/128 (92%) of countries reporting to the WHO had adopted DTG with two NRTIs as first-line therapy.^20^ While NNRTIs are no longer part of first-line therapy, resistance remains of interest as the long-lasting injectable INSTI cabotegravir (CAB) given in combination with the NNRTI rilpivirine (RPV) is being rolled-out throughout sub-Saharan Africa.^21^

Observational studies have so far demonstrated promising results regarding the impact of DTG-based regimens on viral suppression among adults living with HIV. DTG-based regimens have been associated with increased levels of viral suppression among PLHIV initiating treatment and transitioning from a non-DTG regimen in Lesotho^22^, Malawi^23^, South Africa^24^, Tanzania^25^, and in the multi-site AFRICOS cohort (Uganda, Kenya, Tanzania, and Nigeria).^26^ Multi-national World Health Organization data on care-seeking PLHIV, however, do not report significant trends towards increased suppression among those on DTG through 2022^27^. While DTG resistance has been observed in longitudinally-sampled data from individuals failing therapy,^28–31^ only about 5% of all people failing DTG regimens harbor resistance,^27,32^ which is disproportionately concentrated among those with pre-existing NRTI resistance.^23,32^ Clinic-based studies, however, are unable to assess rates of suppression and resistance among all PLHIV, including those not engaged or only transiently engaged in therapy.

Population-based studies that sample individuals regardless of HIV serostatus or engagement in care are needed to address limitations in clinic-based studies and provide a comprehensive assessment of the impact of DTG on population-level virological outcomes. In this study, we used epidemiological and virological data collected from 8,781 people living with HIV aged 15-49 years who participated in a population-based open-cohort study in southern Uganda to assess population- and individual-level viral suppression patterns between 2012 and 2022, concurrent with the scale-up of DTG-based regimens. DTG was first indicated in this setting for men, adolescent boys and women and adolescent girls on contraceptives in 2018 and broadened in 2022 to nearly all PLHIV.^33,34^ HIV deep-sequence data was used to identify ART resistance mutations and evaluate longitudinal resistance trends. This work provides critical insight into the real-world adoption of DTG-based regimens, their efficacy in moving Uganda closer to the UNAIDS 95-95-95 goals for HIV control, and potential implications for future first-line therapy policies in sub-Saharan Africa.

## Methods

### Ethics statement

The Rakai Community Cohort Study is administered by the Rakai Health Sciences Program (RHSP) and has received ethical approval from the Uganda Virus Research Institute’s Research and Ethics Committee (GC/127/08/12/137), the Uganda National Council for Science and Technology (HS540), and the Johns Hopkins Medicine Institutional Review Board (IRB00291604, IRB00217467). Participants provided written informed consent at each survey round. Written assent and written parental consent were obtained for participants less than 18 years of age.

### Study design and participants

The Rakai Community Cohort Study (RCCS) is a population-based open-cohort study conducted every 18-24 months in inland (HIV prevalence ∼14%-17%) and Lake Victoria fishing (∼42%) communities in southern Uganda.^35^ During survey rounds all households in participating communities were censused and consenting residents aged 15-49 were invited to complete a structured sociodemographic, behavioral, and health questionnaire. Participants provided samples for HIV testing,^36^ viral load quantification (Abbott Laboratories real-time m2000), and viral deep-sequencing. Specifically, participants self-reported their age, sex, and whether they have ever accessed HIV care, are aware of their HIV status, or have ever taken HIV ART. Participants were considered pre-treatment at a given survey round if they were seropositive and reported never having received an HIV test result, accessed HIV care, been on long-term medication, or been on ART, and all available viral load measurements taken at that survey or earlier were >1,000 copies/mL.While the analysis period included only the five most recent survey rounds (August 10, 2011 through March 7, 2023), we used self-reported ART exposure from all RCCS survey rounds, beginning on November 5, 1994, to determine participant ART exposure. Throughout, we refer to surveys by the year of the median interview date (appendix 2 p 1).

Viremia was defined as a viral load >1,000 copies/mL. As viral loads in the 2012 survey were only routinely measured among fishing community residents, we imputed missing 2012 viremia status among pre-treatment individuals (*n=*792/2,015, see appendix 1). Observations with missing viral load data in later rounds were dropped from analyses of viremia status (*n*=97/16,885).

Because the RCCS does not ask participants about their drug regimens, we requested data on DTG use among clinic attendees by quarter (quarter 1 2018 through quarter 1 2023) from the 20 most commonly accessed HIV care clinics among RCCS participants in 2022. Among clinics providing data, missing data points were minimal (1/161 of all quarters stratified by sex) and were dropped.

### HIV deep-sequence based drug resistance prediction

HIV deep-sequencing from venous blood samples was performed through the Phylogenetics and Networks for Generalized HIV Epidemics in Africa consortium (PANGEA-HIV) using an overlapping amplicon approach or the validated bait-capture protocol veSEQ-HIV.^37–39^ In addition to RCCS participants, HIV sequence data were also obtained from individuals residing in surrounding areas through complementary research protocols conducted by the Rakai Health Sciences Program. HIV genomes were assembled from quality-filtered reads using Shiver and subtypes were assigned based on the most closely related reference sequence. Complete (≥95%) polymerase sequences were used to calculate Kimura 2-parameter pairwise consensus genetic distances and infer phylogenies using IQ-Tree v.2.2.2.7. (appendix 1).

Drug-resistance mutations (DRMs) were identified using the validated drmSEQ pipeline (appendix 1).^40^ We considered mutations supported by a minimum of 10 PCR-deduplicated reads and at least 5% of reads spanning a respective codon.

Mutations were scored according to the Stanford University HIV Drug Resistance Database v.9.6 and the score sum was used to predict susceptibility to 25 drugs (appendix 2 pp 2-3). We considered a score of ≥30 (intermediate/high-level resistance) as resistant and scores between 10 and 29 as low-level resistance. Resistance was considered indeterminate if <50% of the relevant positions for a drug had <10 reads. We considered a sequence resistant to a given ART class (INSTI, NNRTI, NRTI, PI) if it was assigned intermediate/high-level resistance to any of the drugs within that class. To ensure accurate denominators, class-level resistance was indeterminate if resistance status for any of the drugs within that class was indeterminate.

### Study outcomes and statistical methods

We first estimated the prevalence of HIV, treatment-experienced HIV, and suppressed HIV among all study participants using robust Poisson regression with study round as a categorical predictor and a log-link.^41^ We then estimated the prevalence of ever having been on treatment and of viremia among PLHIV and the prevalence of viremia among treatment-experienced PLHIV (txPLHIV).

The primary outcomes of this study were overall and class-specific prevalence of INSTI, NNRTI, NRTI, and PI resistance among viremic txPLHIV and among pre-treatment PLHIV (ptPLHIV) estimated using robust Poisson regression. To account for missing resistance prediction among some viremic PLHIV, we employed inverse probability weighting (appendix 1). Among viremic txPLHIV, we further estimated the prevalence of multi- and single-class resistance. For some outcomes we estimated the risk ratio associated with age, community type, sex, and viral subtype in both univariate and bivariate (with survey round) models. Finally, we estimated the prevalence of DRMs among viremic txPLHIV and ptPLHIV using participant-visits with successful resistance prediction to all drug classes. We restricted analyses of viremic txPLHIV to 2014 and later due to missing 2012 viral loads.

Because individuals may participate in multiple surveys, prevalence was estimated using generalized estimating equations with geepack v.1.3.11 in R v.4.4.1 using the correlation structure that minimized QIC. For outcomes with <20 outcome events, independence was assumed to ensure convergence. We used an independence correlation structure when estimating the prevalence of individual DRMs to ensure convergence, particularly for rare mutations. Emmeans v.1.10.4 was used to calculate estimated marginal mean values.

Finally, we quantified the individual-level probability of suppression among viremic PLHIV in the preceding survey as a function of survey round and resistance to NNRTI- and NRTI-based regimens. To account for longitudinal changes in treatment initiation and treatment success given initiation we did not stratify this analysis by treatment, but note that suppressed PLHIV are presumed to be treatment-experienced. We accounted for missing data due to drop-outs between surveys and missing viral genomic data using Bayesian post-stratification by survey, age category, community type, and sex (appendix 1).

Unless noted, all analyses were conducted in R with tidyverse v.2.0.0 using ggplot2 v.3.5.1 with patchwork v.1.2.0 and cowplot v.1.1.3 for visualization.

## Results

In six surveys between August 15, 2011 and February 23, 2023 48,914 people, among whom 8,781 (17.95%) were HIV seropositive during the study period, contributed 109,328 participant-visits. Study population demographics remained stable over the study period (appendix 2 p 4), however, the median (IQR) age of participants with HIV increased from 32 (10) to 37 (11) years (appendix 2 p 5).

Over the study, the prevalence of HIV among RCCS participants decreased slightly from 20.5% (95% confidence interval [CI]: 19.9%-21.2%) to 18.1% (95%CI: 17.6%-18.7%, figure 1A and appendix 2 p 6). Concurrent with efforts to improve HIV diagnoses and treatment initiation, including Universal Test and Treat, the prevalence of ever having been on ART (self-reported) among PLHIV increased from 35.4% (95%CI: 33.9%-37.1%) to 94.4% (95%CI: 93.6%-95.2%) over the same period (appendix 2 p 7). Consequently, the prevalence of suppression among PLHIV increased from 57.1% (95% CI: 55.4%-58.8%) to 90.3% (95% CI: 89.2%-91.4%) between 2014 and 2022. The population prevalence of viremic HIV among all participants, which informs the epidemiological risk of HIV acquisition among seronegative participants, decreased 4.6-fold (95%CI: 4.06-5.21) reaching 1.7% (95%CI: 1.6%-2.0%) by 2022. Due to increased treatment, the proportion of people who were treatment-experienced among those with viremia increased 3.14-fold (prevalence ratio [PR] 95%CI: 2.6-3.80), from 13.4% (95%CI: 11.7%-15.3%) in 2014 to 42.1% (95% CI: 36.7%-48.3%) by 2022. Among those people who were treatment experienced, suppression reached 95.4% (95%CI: 94.6%-96.1%) by 2022, an increase from 93.6% (95%CI: 92.8%-94.4%) in 2019, prior to DTG scale-up, largely due to an increase in viral loads ≤ 200 copies/mL (appendix 2 p 8).

**Figure 1:**
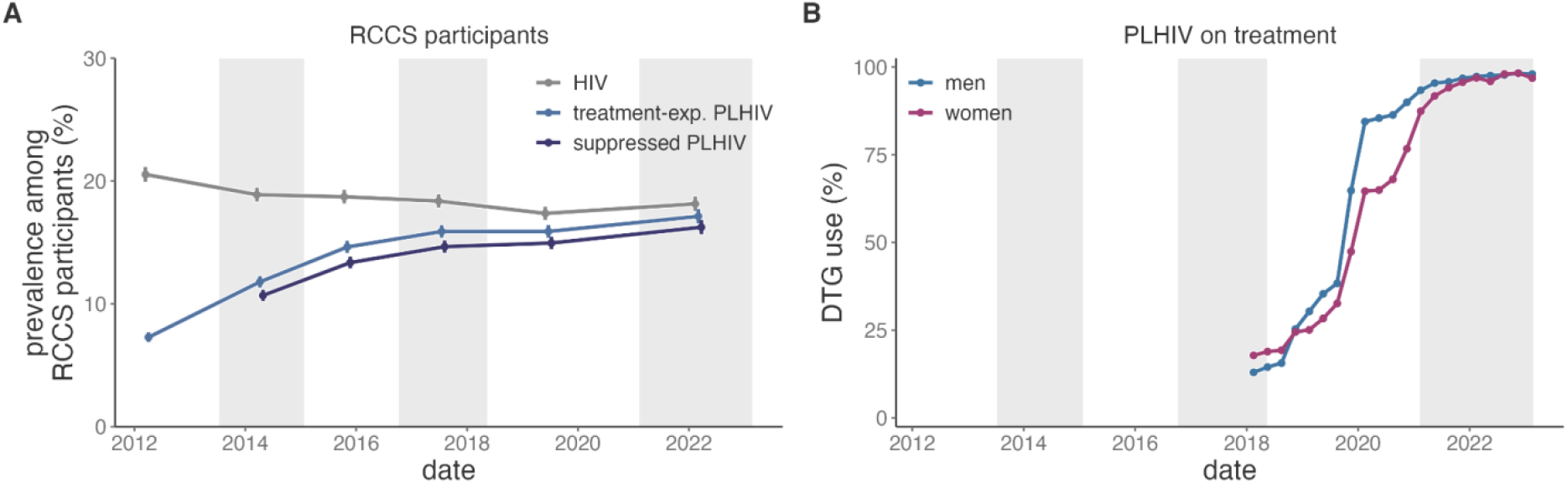
ART scale-up in the Rakai Community Cohort Study (RCCS) 2012-2022. A) Temporal trends in the prevalence of HIV (grey), prevalence of PLHIV who ever report being on ART (light blue), and prevalence of virally suppressed HIV (dark blue). Prevalence estimates were generated using Poisson regression with robust standard errors with survey round as a predictor variable. Generalized estimating equations with correlation structure selection by Quasi Information Criterion value (independence) were used to account for repeat participants across study rounds. B) Proportion of PLHIV on DTG-based regimens at the top 14 clinics serving RCCS participants stratified by sex. Vertical error bars indicate the Wald 95% confidence interval for the mean value but for some estimates do not extend past the size of the point. Shading corresponds to the range of interview dates for alternating RCCS survey rounds.

Overall, 14 clinics that collectively served 74.1% of 2022 RCCS participants reporting ART use provided DTG prescription data (figure 1B and appendix 2 p 9). The proportion of PLHIV at surveyed clinics who were prescribed DTG was 53.6% (13,248/24,735) by the end of 2019 and 97.2% (26,831/27,597) at the end of follow-up (appendix 2 p 10). This increase was slower among women during early 2020 (adjusted *p*-value=0.0002). DTG adoption was consistent across facilities with the exception of Masaka (The AIDS Support Organization) where 3,294/7,211 (45.7%) of PLHIV were prescribed DTG by 2018, Lwanda where DTG use increased rapidly from 0% to 87.2% (135/156 PLHIV) in early 2019, and Kifamba where adoption lagged until 2022 (adjusted *p*-values < 0.0001, appendix 3 p 1).

Among 963 participant-visits contributed by viremic txPLHIV between 2014 and 2022, deep-sequence based drug-resistance prediction was attempted on 936 (97.2%). Among the 348 (37.2%) contributed by participants who initiated treatment within the cohort, median (IQR) time on treatment was 0.90 (0.20) years. Sequencing coverage was sufficient to successfully predict resistance to ≥1 drug class for 783 participant-visits (83.7%) and for all classes for 630 (67.3%). Sequencing success was better at higher viral loads, with the veSEQ-HIV protocol, and for INSTIs compared to other classes (appendix 2 p 11).

Between 2014 and 2022, the prevalence of resistance to any ARTs among viremic txPLHIV decreased significantly from 51.1% (95%CI: 40.7%-64.2%) to 27.9% (95%CI: 21.3%-36.5%, figure 2A and appendix 2 p 12). Although the decline had been occurring since 2015, the biggest drop happened between 2019 and 2022, with a decrease of 4.99% per year (95%CI: -10.10%-0.11% year^-1^, *p*-value=0.0325; figure 2B). Consequently, in 2022, the majority of viremic txPLHIV lacked detectable resistance. These results were robust to a more liberal variant calling threshold (appendix 3 p 2). Resistance among txPLHIV did not vary by age or viral subtype but was slightly more common among participants in inland, as compared to fishing, communities (appendix 2 p 13, adjusted *p*-value=0.053) and women (adjusted *p*-value=0.0001). Over this time frame the vast majority of observed resistance among txPLHIV was NNRTI (2022 prevalence 22.2%, 95%CI: 17.1%-28.9%) and NRTI (12.3%, 95%CI: 7.7%-19.9%). INSTI and PI resistance remained less than 4% over this time frame, with only 2/108 (1.85%) of 2022 txPLHIV exhibiting intermediate/high level DTG resistance (table 1).

**Table 1:**
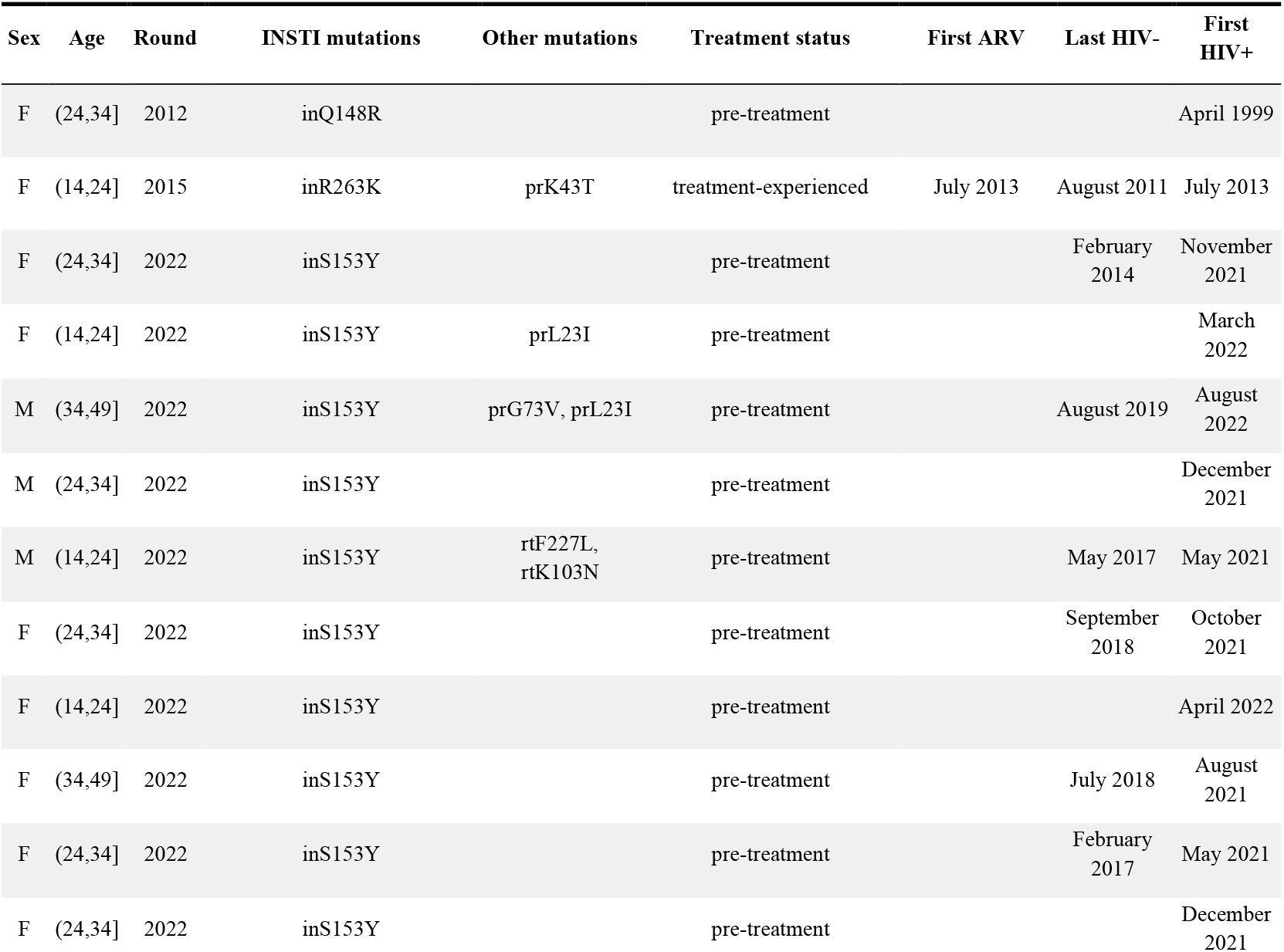
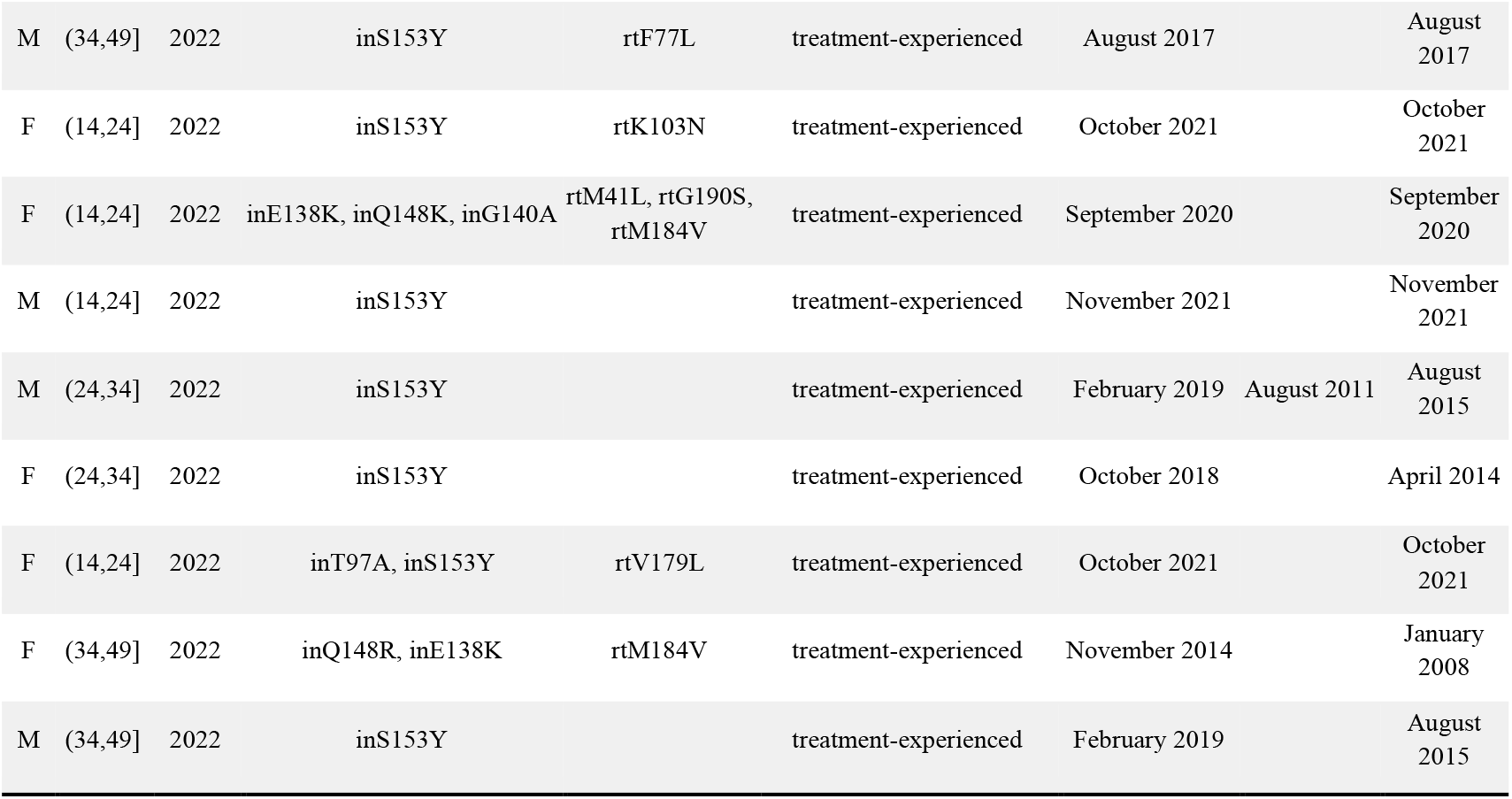
Demographic data on participants with low-, intermediate-, or high-level dolutegravir (DTG) resistance.

**Figure 2:**
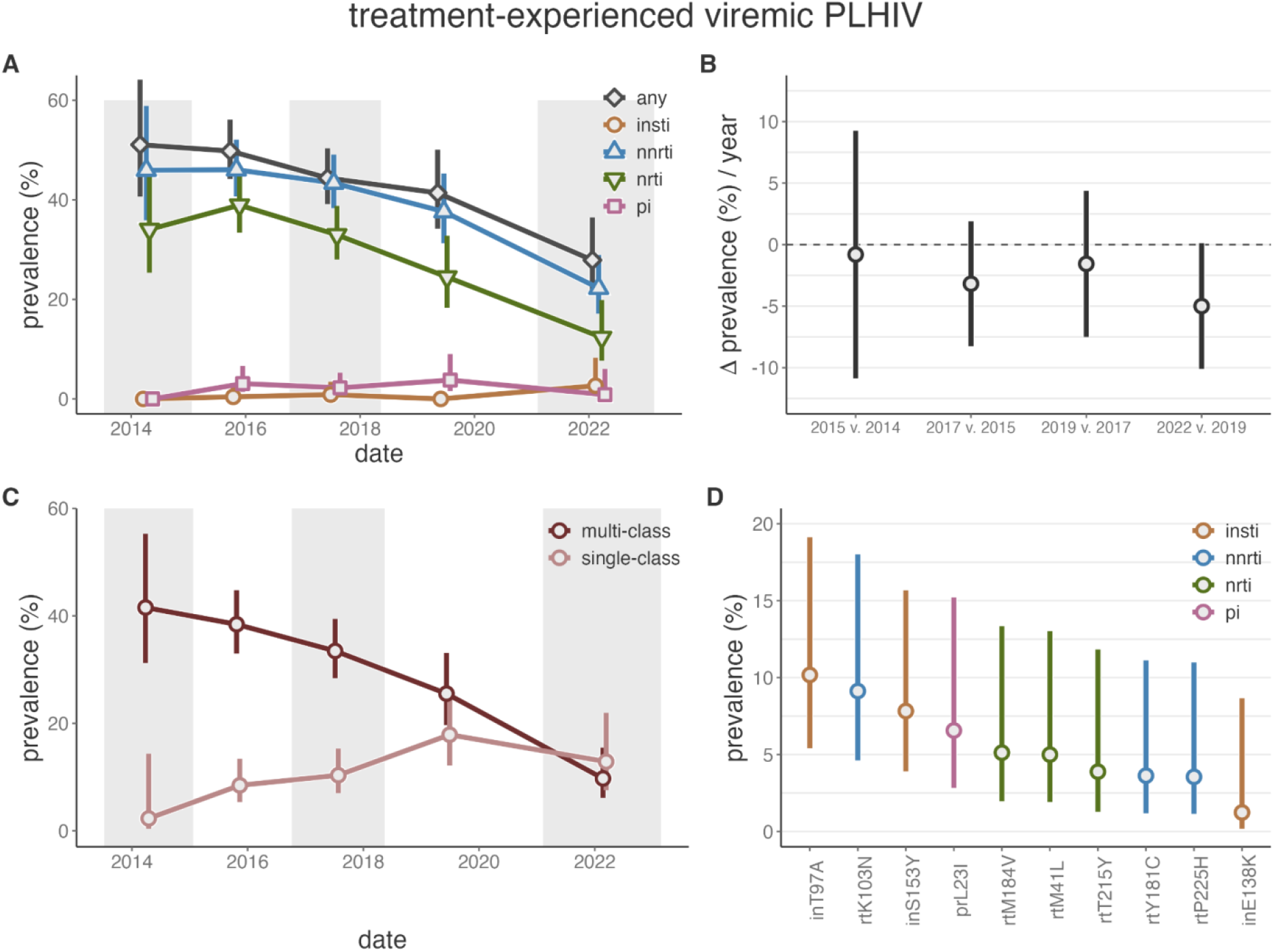
ART resistance among treatment-experienced viremic PLHIV in the Rakai Community Cohort Study. A) Prevalence of resistance to any drug class (grey), integrase strand transfer inhibitors (INSTIs, orange), non-nucleoside reverse transcriptase inhibitors (NNRTIs, blue), nucleoside reverse transcriptase inhibitors (NRTIs, green), and protease inhibitors (PIs, pink) by survey round among treatment-experienced viremic PLHIV. B) Difference between the estimated prevalence of resistance to any drug class in consecutive survey rounds, adjusted for the time between surveys. C) Prevalence of multi-class and single-class resistance among treatment-experienced viremic PLHIV. Multi-class refers to any combination of simultaneous resistance to any of the four drug classes. D) Prevalence of resistance mutations among treatment-experienced viremic PLHIV, colored by the class of drug to which a given mutation confers resistance. Vertical bars extend to the Wald 95% confidence intervals. A,C) Generalized estimating equations with correlation structure selection by Quasi Information Criterion value (Any, NRTI, NNRTI: AR1) or independence for outcome measures with less than 20 events and for all mutations (D) were used to account for repeat participants across study rounds. Shading corresponds to the range of interview dates for alternating RCCS survey rounds.

Resistance among viremic txPLHIV prior to the DTG transition was predominantly multi-class resistance (figure 2C). As the prevalence of NNRTI and NRTI resistance has fallen, the prevalence of multi-class resistance has concurrently fallen significantly (PR 2022 v. 2014 0.23, 95%CI: 0.14-0.4, appendix 2 p 14). Consequently, among viremic txPLHIV in 2022 single-class resistance (12.9%, 95%CI: 7.5%-21.9%) was slightly more common (9.7%, 95% CI: 6.1% - 15.5%).

Among viremic txPLHIV in 2022, the most common resistance mutation was inT97A (10.2%, 95% CI: 5.4%-19.1%), which confers low-level INSTI (e.g. elvitegravir^42^) resistance and has been observed elsewhere in people failing DTG (figure 2D and appendix 2 pp 15-16).^29–31,43^ Another mutation that confers low-level (<three-fold) DTG resistance,^44^ inS153Y, was observed in 7/108 (6.48%), of viremix txPLHIV in 2022, despite not being observed earlier (2022 prevalence: 7.8%, 95%CI: 3.9%-15.7%). The majority of NNRTI and NRTI resistance in this population was attributable to rtK103N (9.1%, 95%CI: 4.6%-18.0%, high-level EFV and nevirapine [NVP] resistance^45^), rtM184V (5.1%, 95% CI: 2% - 13.3%], high-level 3TC and emtricitabine [FTC] resistance^46^), and rtM41L (5% [95% CI: 1.9% - 13%], a Thymidine Analog Mutation^47,48^). The prevalence of these mutations decreased between 2014 and 2022 (PR: 0.37, 95%CI: 0.15-08.9; 0.11, 95%CI: 0.04-0.3; 0.45%, 95%CI: 0.12 - 1.66).

Of the two participants with high/intermediate level DTG resistance, one harbored inQ148R with inE138K, conferring about three-fold reduced DTG susceptibility,^49^ as well as rtM184V (NRTI, table 1). The other harbored multi-class INSTI/NRTI/NNRTI resistance attributable to inE138K, inG140A, and Q148R along with rtM41L, rtM184V, and rtG190S.^50,51^ Both individuals had multi-class NNRTI/NRTI resistance before the DTG transition.

Between 2012 and 2022, ptPLHIV contributed 4,981 participant-visits among which deep-sequence based drug-resistance prediction was attempted on 4,924 (98.9%, appendix 2 p 17). Drug-resistance prediction was successful for ≥1 drug class for 3,918 (79.6%) and for all for 2,461 (50%). As above, sequencing success was better at higher viral load and with VeSeq-HIV.

Over the study-period, we observed a 2.72-fold (95%CI: 1.67-4.44) increase in NNRTI resistance among ptPLHIV (figure 3A, appendix 2 p 18). Between 2019 and 2022 alone NNRTI resistance among ptPLHIV increased from 11.5% (95%CI: 8.6%-15.5%) to 14.8% (95%CI: 9.7%-22.7%). NNRTI resistance was slightly more common among women (adjusted *p*-value=0.021, appendix 2 p 19). Intermediate/high-level resistance to NRTIs/INSTIs/PIs remained stable and below 2.5%. Intermediate/high-level DTG resistance was not identified among ptPLHIV.

**Figure 3:**
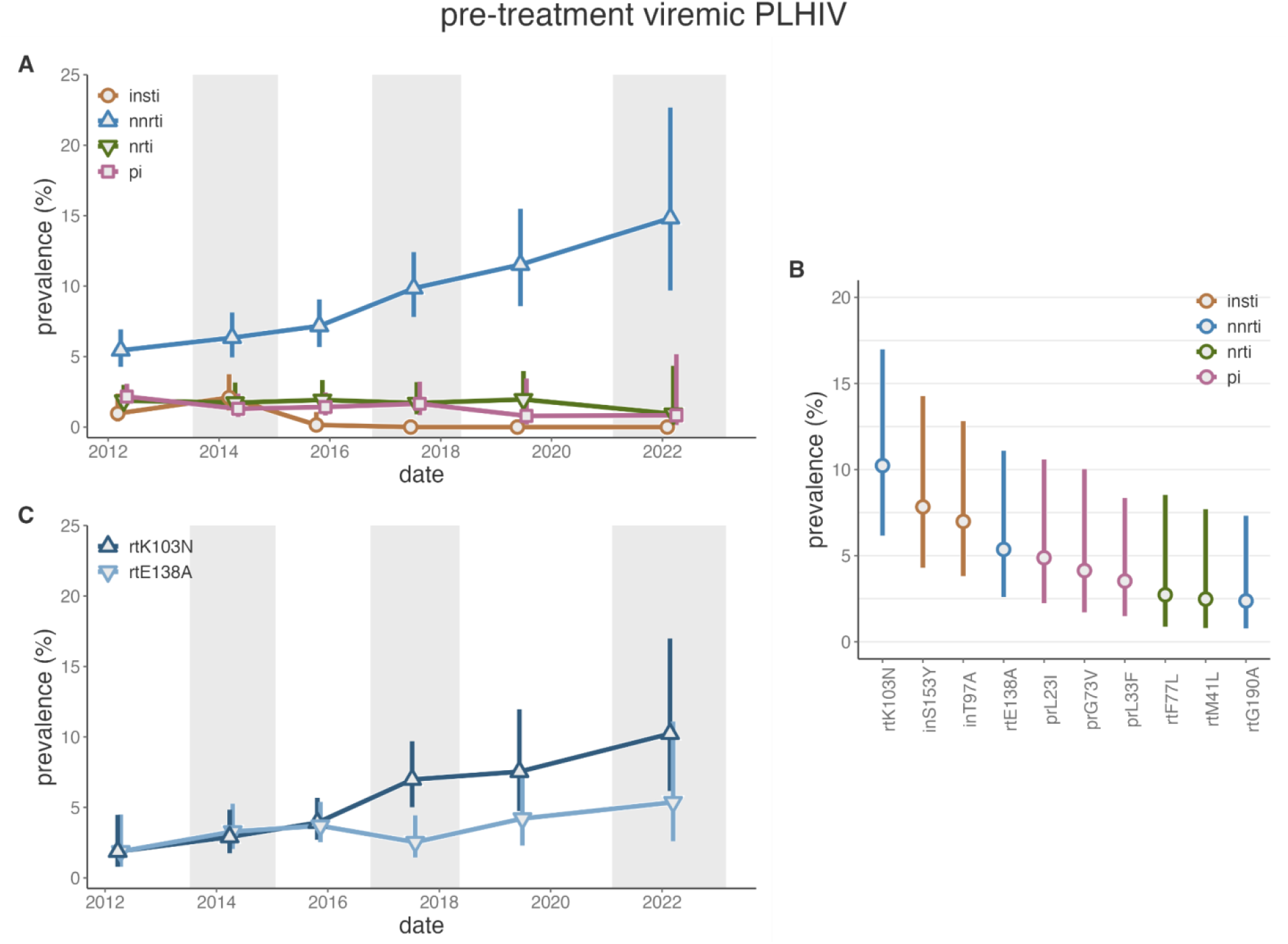
ART resistance among pre-treatment viremic PLHIV in the Rakai Community Cohort Study. A) Prevalence of resistance to integrase strand transfer inhibitors (INSTIs, orange), non-nucleoside reverse transcriptase inhibitors (NNRTIs, blue), nucleoside reverse transcriptase inhibitors (NRTIs, green), and protease inhibitors (PIs, pink) by survey round among treatment-experienced viremic PLHIV. Generalized estimating equations with correlation structure selection by Quasi Information Criterion value (NRTI, PI: AR1, NRTI: exchangeable) or independence for outcome measures with less than 20 events. B) Prevalence of resistance mutations among pre-treatment viremic PLHIV, colored by the class of drug to which a given mutation confers resistance. C) Prevalence of rtK103N and rtE138A mutations by survey round among pre-treatment viremic PLHIV. Bars extend to the Wald 95% confidence intervals. Bars extend to the Wald 95% confidence intervals. Shading corresponds to the range of interview dates for alternating RCCS survey rounds.

NNRTI resistance was predominantly attributed to rtK103N (2022 prevalence: 10.2%, 95%CI: 6.2%-17%, figure 3B,C and appendix 2 pp 20-21) and rtE138A (5.4%, 95%CI: 2.6%-11.1%). The increased prevalence of NNRTI resistance was largely driven by rtK103N which increased 5.49-fold (95%CI: 2-15.06) between 2012 and 2022 whereas rtE138A increased modestly (PR: 2.86, 95%CI: 0.92-8.92). While not solely conferring intermediate/high-level INSTI resistance, we observed inT97A and inS153Y each in 10/170 (5.88%) of pre-treatment PLHIV in 2022 (2022 prevalence: 7%, 95%CI: 3.8%-12.8% and 7.8%, 95%CI: 4.3%-14.3%). While inT97A remained stable over the study period (PR v. 2012: 1.09, 95%CI: 0.51-2.34), inS153Y was not observed prior to 2022, consistent with viremic txPLHIV (table 1).

To assess longitudinal dynamics of suppression among those with and without resistance we used paired individual viral load measurements in consecutive surveys. Between 2015 and 2022, the probability of achieving viral load suppression among all viremic PLHIV in the previous survey increased modestly by 1.38-fold (95% highest posterior density [HPD] 1.23-1.52, appendix 3 p 3). In 2015, viral suppression was slightly less frequent among those with NNRTI and NRTI resistance (RR v. those without resistance: 0.72, 95%HPD: 0.42-1.01 and 0.65, 95%HPD: 0.28-1.02, figure 4). However, over the study period, suppression among those with resistance increased significantly and by 2022 suppression levels did not vary by NNRTI or NRTI resistance (RR: 1.14, 95%HPD: 0.96-1.32, and 1.13, 95%HPD: 0.88-1.35). Notably, this trend was observed throughout the study period, suggestive of improved clinical management during the expansion of HIV treatment programs and potentially increased suppression due to the DTG transition in the 2022 survey.

**Figure 4:**
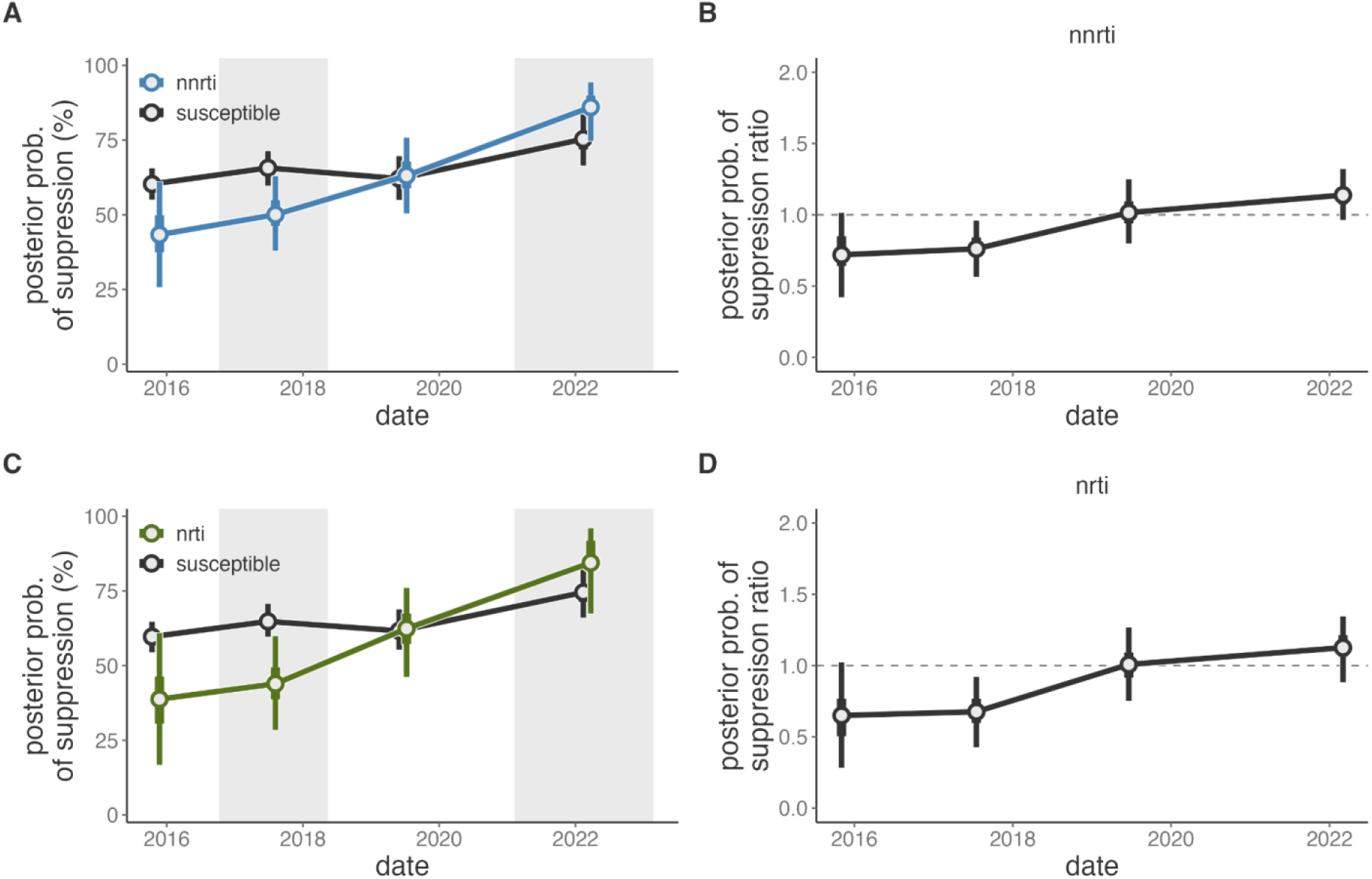
Probability of viral load suppression among Rakai Community Cohort Study participants who were viremic in the proceeding survey round. A) Probability of viral load suppression among PLHIV who participated in the RCCS and were viremic in the preceding survey round, stratified by NNRTI resistance (blue) or susceptibility (black). B) Posterior ratio of the probability of suppression among those with NNRTI resistance compared to those without in each survey round. C) Probability of viral load suppression among PLHIV who participated in the RCCS and were viremic in the preceding survey round, stratified by NRTI resistance (green) or susceptibility (black). B) Risk ratio of the probability of suppression among those with NRTI resistance compared to those without in each survey round. Median value of the posterior distribution plotted as the central estimate and bars extend to the 50% and 95% highest posterior density. Estimates are plotted at the median date of the follow-up survey. Shading corresponds to the range of interview dates for alternating RCCS survey rounds.

As noted, inS153Y was first observed in this setting after the transition to DTG-based regimens. To assess the evolutionary dynamics giving rise to the emergence of this mutation, we integrated RCCS data with available HIV deep-sequence data from other PLHIV in the region (Methods). inS153Y was observed in 23/307 (7.5%) of all deep-sequenced samples collected after the beginning of the 2022 survey. Of these, 11 were living with HIV subtype A1, six with subtype D, and six with recombinant subtypes. The mutation was universally observed as a minor variant, present in 5-10% of sequence reads and there was no obvious pattern of it co-occurring with other resistance mutations (appendix 3 pp 4-5). Five individuals with inS153Y (21.74%) were linked to another case of inS153Y at a genetic distance less than 0.065 substitutions/site, the 99th percentile of pairwise distances between viremic PLHIV without inS153Y sampled over the same epoch (figure 5). We observed two inS153Y genetic clusters, one of size three (subtype D, two females and a male) and one of size two (subtype A1, female and a male), indicative of multiple instances of linkage through recent transmission.

**Figure 5:**
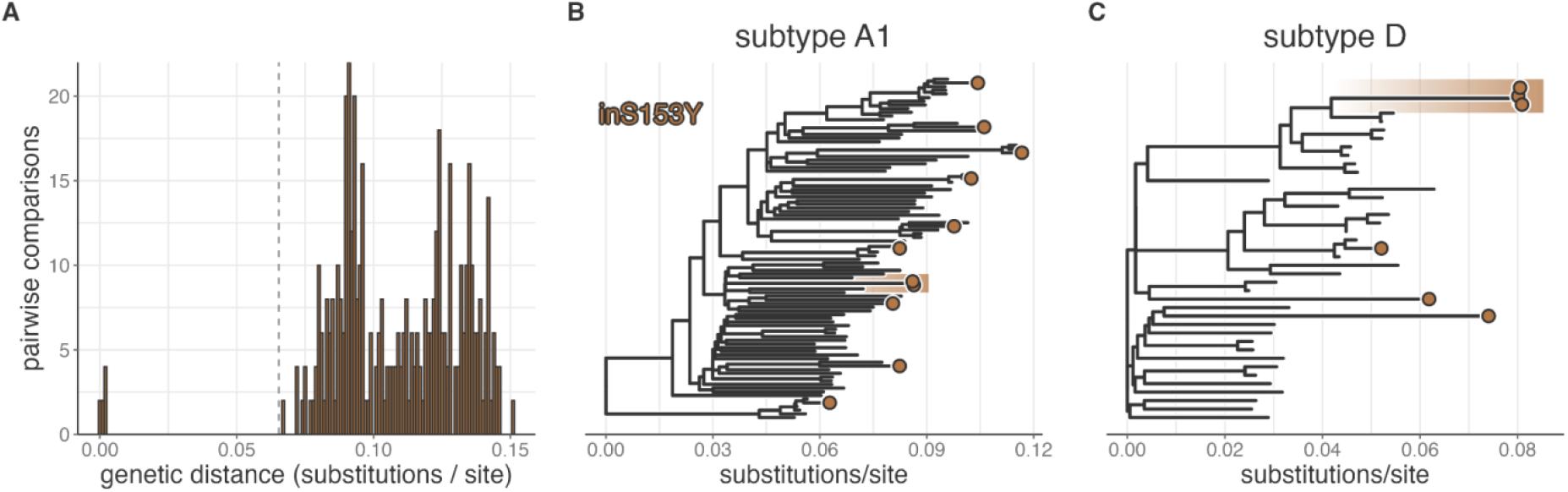
Genetic clustering of Rakai residents living with viremic HIV with the inS153Y mutation. A) Pairwise viral genetic distance between Rakai residents with the inS153Y mutation, all of which were sampled after February 8, 2021. Dashed vertical line indicates the 99th percentile of pairwise genetic distance between people living with HIV without the inS153Y mutation sampled between February 8, 2021 and the end of the study period, September 7, 2023. B) Phylogenetic tree of subtype A1 HIV sequences from PLHIV with the inS153Y mutation (orange tips) and the 10 most closely related Rakai HIV sequences for each. C) Phylogenetic tree of subtype D HIV sequences from PLHIV with the inS153Y mutation (orange tips) and the 10 most closely related Rakai HIV sequences for each. In B and C clustered tips as identified in A are highlighted with orange background.

## Discussion

Here, we quantified HIV treatment utilization, viral load suppression, and ART resistance throughout a period of intensive scale-up of treatment and prevention efforts, including the transition to DTG-based first line regimens. Population-level viral suppression significantly increased over the study period and by 2022 over 90% of PLHIV were suppressed. Among viremic txPLHIV we observed an almost halving in the overall prevalence of resistance, indicative of increased suppression among individuals with resistance. Indeed, individual-level viral load trajectories revealed that suppression among viremic people with a history of NNRTI and NRTI resistance increased roughly two-fold over the study. Despite the DTG-transition, pre-treatment NNRTI resistance continued to increase. We observed intermediate/high-level DTG resistance among only two individuals, both treatment-experienced, in the years immediately following the transition to DTG regimens. However, we observed the emergence and potential transmission of a low-level INSTI resistance mutation, inS153Y, among both pre-treatment and treatment-experienced people with HIV as a minor variant concurrent with DTG scale-up. These results provide critical insights into the dynamics of HIV treatment and suppression in an East-African setting with high-HIV burden with important implications for efforts to support reaching milestones such as the UNAIDS 95-95-95 targets.

These findings from a population-based study corroborate clinic-based studies on the impact of DTG on viral suppression. In South Africa, for example, DTG initiators were slightly more likely to achieve suppression after 12-months (83% v. 81%).^24^ Among PLHIV on treatment, those who transitioned to DTG-regimens were more likely to maintain suppression (94.3% v. 82.1%) in the AFRICOS (Kenya, Nigeria, Tanzania, and Uganda) cohort^26^ and overall suppression rate among DTG-initiators was marginally, yet significantly, higher in South Africa (90.5% v. 89.7%).^24^ People initiating or transitioning to DTG regimens in South Africa were also 1.09- and 1.03-times more likely to be retained in care at 12 months. Here, we build upon these findings by showing that viral load suppression among PLHIV increased significantly, from 86.1% (2019) to 90.3% (2022) following the DTG transition. This increase followed a trend ongoing since 2014 and likely reflects the combined impact of DTG and programmatic changes to support treatment-initiation and sustained adherence.^52–55^

Consistent with prior work we observed extremely limited evidence of intermediate/high-level DTG-resistance among viremic people. Of the two individuals with DTG resistance both had pre-existing NRTI resistance, consistent with reports that NRTI resistance increases the risk of acquired DTG resistance.^23,32^ Despite limited intermediate/high-level INSTI resistance, the low-level (<three-fold^44^) resistance mutation inS153Y was observed among txPLHIV and ptPLHIV in 2022, despite not being observed earlier. Concerningly, a small subset of PLHIV with inS153Y may be linked by recent transmission based on HIV genetic clustering. This pattern could be indicative of recent transmission from unsampled individuals failing DTG followed by reversion of deleterious mutations conferring high-level DTG resistance. Additional analyses of the viral evolutionary dynamics associated with harboring inS153Y are needed to further validate this hypothesis. More generally, observing inS153Y in this setting soon after the DTG transition highlights the value of population-based genomic surveillance for identifying emerging viral mutations following new treatment regimens. Further, the continued increase in the prevalence of rtE138A (RPV resistance) among ptPLHIV requires monitoring in light of efforts to roll-out long-lasting injectable CAB/RPV.^56^ This mutation was observed in 2/4 individuals failing CAB/RPV at 96-weeks in the Cabotegravir and Rilpivirine Efficacy and Safety Trial.^57^ Pre-treatment resistance testing, not currently routine in sub-Saharan Africa, may be necessary to ensure individuals with rtE138A achieve durable suppression on CAB/RPV. Continued monitoring of INSTI-resistance mutations among people failing therapy and on a population level, is needed to ensure that continued DTG utilization does not select for the emergence of resistance mutations among PLHIV on therapy.

NNRTI and NRTI resistance among viremic txPLHIV declined throughout the study period, a trend that accelerated during the DTG transition. This is suggestive of increased suppression among those with resistance, as indicated by our analyses, and reductions in acquired resistance due to e.g. intermittent treatment adherence. The acceleration of this trend is consistent with DTG-driven suppression among individuals with resistance. In 2022, the majority (72.1%) of viremic txPLHIV did not have detectable intermediate/high-level ART resistance and 97.3% were susceptible to DTG. As NRTI resistance rapidly reverts during treatment interruption,^58^ treatment-experienced NRTI resistance may be a marker of transient care engagement and risk of DTG failure. On the contrary, viremic txPLHIV without resistance may be disengaged from care entirely. In this study, this was more likely among men, suggestive of worse ART adherence among men.^59^ As 42.1% of viremic people are treatment-experienced, re-engaging this population in care is critical in reducing the population prevalence of viremia.

While we assess DTG utilization among regional clinics, we lack universal data on individual-level regimens and cannot estimate a direct effect of DTG on viral suppression. Similarly, drug presence assays and phenotypic resistance testing are not routinely conducted in the RCCS and we therefore cannot disentangle the role of adherence and resistance in viremia among txPLHIV. Throughout, we rely on self-reported treatment status, possibly leading to misclassification particularly among ptPLHIV. We suspect misclassification to be minimal given the divergent resistance motifs among txPLHIV and ptPLHIV and minimal (11%) ART use among pre-treatment RCCS participants in previous work.^60^

Together, these results provide an encouraging view of the HIV epidemic in this East African setting. At the end of follow-up in early 2023, the most significant barriers to widespread suppression on DTG-based regimens appear to be delays in treatment initiation and failure to achieve persistent adherence. The risk of DTG resistance, thus far, appears to be minimal, particularly among ptPLHIV. The rapid emergence of inS153Y suggests the need for ongoing viral genomic surveillance to maintain the long-term DTG efficacy. This highlights the fact that investments in developing ART-regimens and making them widely available has been instrumental in achieving significant reductions in viremia and therefore HIV-related disease^1^ and transmission.^61^ Resistance has not, to date, eroded these population-scale impacts. However, as HIV care is suppressive, and not curative, any reduction in access to treatment threatens to rapidly erode many of the positive impacts of treatment scale-up.^62–64^ Further, as intermittent adherence to INSTI-based regimens is associated with a significantly increased rate of acquired resistance,^65^ treatment interruptions may lead to wide-scale emergence of resistance against a highly effective ART regimen, thwarting major public health and research investments. This would have devastating consequences for the health of people living in high-HIV burden communities as well as people globally given the international nature of HIV transmission dynamics.^66^

## Data Availability

All code and processed de-identified data (with the exception of genetic linkage data) needed to reproduce the results of this study are available at https://github.com/m-a-martin/rccs_hiv_dtg_resistance. A subset of HIV consensus sequences are available from Zenodo (https://doi.org/10.5281/zenodo.10075814) and the PANGEA-HIV (https://github.com/PANGEA-HIV/PANGEA-Sequences) sequence repository under the CC-BY-4.0 license. Due to privacy and ethical reasons and in alignment with UNAIDS ethical guidelines, HIV deep-sequence data can be requested from PANGEA-HIV under a managed access policy. More information on data requests can be accessed through PANGEA-HIV (https://www.pangea-hiv.org/join-us). Additional RCCS data can be requested from the Rakai Health Sciences Program (RHSP) under a managed access policy upon request from.

https://github.com/m-a-martin/rccs_hiv_dtg_resistance/tree/main

https://doi.org/10.5281/zenodo.10075814

https://github.com/PANGEA-HIV/PANGEA-Sequences

## Acknowledgements

This work was supported by the United States of America National Institutes of Health (NIH) National Institute of Allergy and Infectious Diseases (NIAID) (U01AI075115, R01AI087409, U01AI100031, R01AI110324, R01AI114438, K25AI114461, R01AI123002, K01AI125086, R01AI128779, R01AI143333, R21AI145682, R01AI155080, ZIAAI001040), NIH National Institute of Child Health and Development (R01HD050180, R01HD070769, R01HD091003), NIH National Heart, Lung, and Blood Institute (R01HL152813), the Fogarty International Center (D43TW009578, D43TW010557), the Johns Hopkins University Center for AIDS Research (P30AI094189), the Bill & Melinda Gates Foundation (OPP1084362, INV-007573, INV-035619, INV-060259, INV-075093), and the U.S. President’s Emergency Plan for AIDS Relief through the Centers for Disease Control and Prevention (NU2GGH000817). S.J.R., T.C.Q., and O.L. are employed by the NIH. This research was supported [in part] by the Intramural Research Program of the National Institutes of Health (NIH). The contributions of the NIH author(s) were made as part of their official duties as NIH federal employees, are in compliance with agency policy requirements, and are considered Works of the United States Government. However, the findings and conclusions presented in this paper are those of the author(s) and do not necessarily reflect the views of the NIH or the U.S. Department of Health and Human Services.

## Data sharing

All code and processed de-identified data (with the exception of genetic linkage data) needed to reproduce the results of this study are available at https://github.com/m-a-martin/rccs_hiv_dtg_resistance. A subset of HIV consensus sequences are available from Zenodo (https://doi.org/10.5281/zenodo.10075814) and the PANGEA-HIV (https://github.com/PANGEA-HIV/PANGEA-Sequences) sequence repository under the CC-BY-4.0 license. Due to privacy and ethical reasons and in alignment with UNAIDS ethical guidelines, HIV deep-sequence data can be requested from PANGEA-HIV under a managed access policy. More information on data requests can be accessed through PANGEA-HIV (https://www.pangea-hiv.org/join-us,pangea.data.enquiries@ndm.ox.ac.uk). Additional RCCS data can be requested from the Rakai Health Sciences Program (RHSP) under a managed access policy upon request from.

