## Appendix 2 for "Patterns of HIV-1 viral load suppression and drug resistance during the dolutegravir transition: a population-based longitudinal study"

**Supplementary Table 1: Rakai Community Cohort Study survey dates**

| Round | Survey start date | Survey mid date | Survey end date |
|---|---|---|---|
| 1 | 1994-11-05 | 1995-03-20 | 1995-07-18 |
| 2 | 1995-01-12 | 1996-01-10 | 1996-05-01 |
| 3 | 1996-01-12 | 1996-11-20 | 1997-04-23 |
| 4 | 1997-06-06 | 1997-11-21 | 1998-05-12 |
| 5 | 1997-08-05 | 1998-08-06 | 1999-03-15 |
| 6 | 1999-04-06 | 1999-09-07 | 2000-02-09 |
| 7 | 2000-03-20 | 2000-08-08 | 2001-02-19 |
| 8 | 2001-04-03 | 2001-10-23 | 2002-05-31 |
| 9 | 2002-07-15 | 2003-01-28 | 2003-08-01 |
| 10 | 2003-09-26 | 2004-04-23 | 2004-11-23 |
| 11 | 2005-01-13 | 2005-10-13 | 2006-06-30 |
| 12 | 2006-03-31 | 2007-06-28 | 2008-04-24 |
| 13 | 2008-06-17 | 2009-03-17 | 2009-12-04 |
| 14 | 2010-01-18 | 2010-09-24 | 2`011-06-21 |
| 15 | 2011-08-10 | 2012-04-02 | 2013-05-29 |
| 16 | 2013-07-08 | 2014-04-07 | 2015-01-28 |
| 17 | 2015-02-23 | 2015-11-03 | 2016-09-02 |
| 18 | 2016-10-03 | 2017-07-17 | 2018-05-21 |
| 19 | 2018-06-19 | 2019-06-20 | 2020-11-04 |
| 20 | 2021-02-01 | 2022-03-04 | 2023-03-07 |

**Supplementary Table 2: Antiretroviral therapy to which resistance was predicted**

| Generic name | Abbreviation | Class |
|---|---|---|
| bictegravir | BIC | INSTI |
| cabotegravir | CAB | INSTI |
| dolutegravir | DTG | INSTI |
| elvitegravir | EVG | INSTI |
| raltegravir | RAL | INSTI |
| doravirine | DOR | NNRTI |
| efavirenz | EFV | NNRTI |
| Etravirine | ETR | NNRTI |
| nevirapine | NVP | NNRTI |
| rilpivirine | RPV | NNRTI |
| abacavir | ABC | NRTI |
| didanosine | DDI | NRTI |
| emtricitabine | FTC | NRTI |
| lamivudine | 3TC | NRTI |
| stavudine | D4T | NRTI |
| tenofovir | TDF | NRTI |
| zidovudine | AZT | NRTI |
| atazanavir | ATV | PI |
| darunavir | DRV | PI |
| fosamprenavir | FPV | PI |
| indinavir | IDV | PI |
| lopinavir | LPV | PI |
| nelfinavir | NFV | PI |
| saquinavir | SQV | PI |
| tipranavir | TPV | PI |

**Supplementary Table 3: Recommended ART regimen among RCCS participants by year**

| Year | First line regimen | Second line regimen |
|---|---|---|
| 2012 | **Preferred** | **Preferred** |
| | TDF/3TC + NVP | AZT + 3TC + ATV/r |
| | TDF/3TC + EFV | AZT + 3TC + LPV/r |
| | **Alternative** | **Alternative** |
| | AZT/3TC + NVP | TDF + 3TC + ATV/r |
| | AZT/3TC + EFV | TDF + 3TC + LPV/r |
| 2016 | **Preferred** | **Preferred** |
| | TDF+3TC+EFV | 2 NRTIs and ritonavir-boosted atazanavir (ATV/r) |
| | | e.g. |
| | **Alternative (only if EFV is contraindicated)** | After failing on TDF + 3TC or ABC+3TC based regimen: AZT+3TC / ATV/r |
| | TDF+3TC+DTG | |
| | | After failing on AZT+3TC based regimen: TDF + 3TC/ ATV/r |
| 2018 | **Preferred** | **Preferred** |
| | TDF+3TC+DTG | AZT+3TC+ATV/r |
| | **Alternative** | **Alternative** |
| | TDF+3TC+EFV, ABC+3TC+DTG | AZT+3TC+LPV/r |
| | | AZT+3TC+DTG |
| | **Adult women and adolescent girls of childbearing potential who are pregnant, intend to get pregnant or not on effective contraception** | |
| | TDF+3TC+EFV | |
| | **Alternative** | |
| | TDF+3TC+ATVr or ABC+3TC+EFV | |
| 2022 | **Preferred** | **Preferred** |
| | TDF + 3TC + DTG | AZT+3TC+ATV/r |
| | | TDF+3TC+ATV/r |
| | **Alternatives** | |
| | TDF + 3TC + EFV400 | **Alternative** |
| | ABC + 3TC +DTG | AZT+3TC+LPV/r |
| | ABC +3TC +EFV400 | TDF+3TC+LPV/r |
| | TDF +3TC + ATV/r or ABC | |
| | 3TC + ATV/r | |

**Supplementary Table 4: Demographics of RCCS participants by survey round.**

| | | Survey Round | | | | | |
|---|---|---|---|---|---|---|---|
| | | 2012 | 2014 | 2015 | 2017 | 2019 | 2022 |
| Overall | | 17,034 | 17,979 | 19,317 | 19,775 | 19,276 | 15,947 |
| Age (median [IQR]) | | 28 [13] | 29 [15] | 29 [14] | 29 [16] | 29 [17] | 30 [18] |
| Age category | | | | | | | |
| | (14,24] | 5,977 (35.1%) | 6,404 (35.6%) | 6,813 (35.3%) | 6,968 (35.2%) | 6,679 (34.6%) | 5,468 (34.3%) |
| | (24,34] | 6,394 (37.5%) | 6,282 (34.9%) | 6,411 (33.2%) | 6,358 (32.2%) | 5,913 (30.7%) | 4,607 (28.9%) |
| | (34,49] | 4,663 (27.4%) | 5,293 (29.4%) | 6,093 (31.5%) | 6,449 (32.6%) | 6,684 (34.7%) | 5,872 (36.8%) |
| Community type | | | | | | | |
| | Agrarian | 7,586 (44.5%) | 7,947 (44.2%) | 8,565 (44.3%) | 8,488 (42.9%) | 8,589 (44.6%) | 5,747 (36%) |
| | Fishing | 3,851 (22.6%) | 3,923 (21.8%) | 4,234 (21.9%) | 4,748 (24%) | 4,058 (21.1%) | 4,343 (27.2%) |
| | Trading | 5,597 (32.9%) | 6,109 (34%) | 6,518 (33.7%) | 6,539 (33.1%) | 6,629 (34.4%) | 5,857 (36.7%) |
| Sex | | | | | | | |
| | F | 9,194 (54%) | 9,669 (53.8%) | 10,446 (54.1%) | 10,534 (53.3%) | 10,415 (54%) | 8,537 (53.5%) |
| | M | 7,840 (46%) | 8,310 (46.2%) | 8,871 (45.9%) | 9,241 (46.7%) | 8,861 (46%) | 7,410 (46.5%) |

Percentages represent the proportion of total participant-visits in each survey round belonging to each sub-category.

**Supplementary Table 5: Demographics of RCCS participants living with HIV by survey round.**

| | | Survey Round | | | | | |
|---|---|---|---|---|---|---|---|
| | | **2012** | **2014** | **2015** | **2017** | **2019** | **2022** |
| Overall | | 3,498 | 3,396 | 3,616 | 3,633 | 3,347 | 2,893 |
| Age (median [IQR]) | | 32 [10] | 33 [12] | 34 [11] | 34 [11] | 36 [12] | 37 [11] |
| Age category | | | | | | | |
| | (14,24] | 532 (15.2%) | 481 (14.2%) | 450 (12.4%) | 366 (10.1%) | 267 (8%) | 199 (6.9%) |
| | (24,34] | 1,663 (47.5%) | 1,471 (43.3%) | 1,483 (41%) | 1,451 (39.9%) | 1,211 (36.2%) | 931 (32.2%) |
| | (34,49] | 1,303 (37.2%) | 1,444 (42.5%) | 1,683 (46.5%) | 1,816 (50%) | 1,869 (55.8%) | 1,763 (60.9%) |
| Community type | | | | | | | |
| | Agrarian | 1,049 (30%) | 1,043 (30.7%) | 1,099 (30.4%) | 1,031 (28.4%) | 1,027 (30.7%) | 650 (22.5%) |
| | Fishing | 1,598 (45.7%) | 1,522 (44.8%) | 1,600 (44.2%) | 1,737 (47.8%) | 1,503 (44.9%) | 1,534 (53%) |
| | Trading | 851 (24.3%) | 831 (24.5%) | 917 (25.4%) | 865 (23.8%) | 817 (24.4%) | 709 (24.5%) |
| Sex | | | | | | | |
| | F | 2,171 (62.1%) | 2,111 (62.2%) | 2,297 (63.5%) | 2,240 (61.7%) | 2,142 (64%) | 1,848 (63.9%) |
| | M | 1,327 (37.9%) | 1,285 (37.8%) | 1,319 (36.5%) | 1,393 (38.3%) | 1,205 (36%) | 1,045 (36.1%) |

Percentages represent the proportion of total participant-visits in each survey round belonging to each sub-category.

**Supplementary Table 6: Prevalence of HIV, treatment-experienced HIV, and viremic HIV among all RCCS participants.**

| | | HIV | | | | Treatment-experienced PLHIV | | | | | Viremic HIV | | | | |
|---|---|---|---|---|---|---|---|---|---|---|---|---|---|---|---|
| **Survey round** | ***n*** | **Obs (%)** | **Prev. (95% CI)** | **Prev. ratio (95% CI)** | ***p*-value** | **n** | **Obs (%)** | **Prev. (95% CI)** | **Prev. ratio (95% CI)** | ***p*-value** | **n** | **Obs (%)** | **Prev. (95% CI)** | **Prev. ratio (95% CI)** | ***p*-value** |
| 2012 | 17,034 | 3,498 (20.5%) | 20.5 (19.9, 21.2) | ref | ref | 17,034 | 1,240 (7.3%) | 7.3 (6.9, 7.7) | ref | ref | | | | | |
| 2014 | 17,979 | 3,396 (18.9%) | 18.9 (18.3, 19.5) | 0.92 (0.89, 0.95) | <0.0001 | 17,979 | 2,120 (11.8%) | 11.8 (11.3, 12.3) | 1.62 (1.54, 1.7) | <0.0001 | 17,942 | 1,442 (8%) | 8 (7.6, 8.4) | ref | ref |
| 2015 | 19,317 | 3,616 (18.7%) | 18.7 (18.2, 19.3) | 0.91 (0.88, 0.94) | <0.0001 | 19,317 | 2,826 (14.6%) | 14.6 (14.1, 15.1) | 2.01 (1.91, 2.12) | <0.0001 | 19,311 | 1,030 (5.3%) | 5.3 (5, 5.7) | 0.66 (0.62, 0.71) | <0.0001 |
| 2017 | 19,775 | 3,633 (18.4%) | 18.4 (17.8, 18.9) | 0.89 (0.87, 0.92) | <0.0001 | 19,775 | 3,143 (15.9%) | 15.9 (15.4, 16.4) | 2.18 (2.07, 2.31) | <0.0001 | 19,765 | 726 (3.7%) | 3.7 (3.4, 3.9) | 0.46 (0.42, 0.5) | <0.0001 |
| 2019 | 19,276 | 3,347 (17.4%) | 17.4 (16.8, 17.9) | 0.85 (0.82, 0.88) | <0.0001 | 19,276 | 3,064 (15.9%) | 15.9 (15.4, 16.4) | 2.18 (2.06, 2.31) | <0.0001 | 19,262 | 453 (2.4%) | 2.4 (2.1, 2.6) | 0.29 (0.26, 0.32) | <0.0001 |
| 2022 | 15,947 | 2,893 (18.1%) | 18.1 (17.6, 18.7) | 0.88 (0.85, 0.92) | <0.0001 | 15,947 | 2,731 (17.1%) | 17.1 (16.6, 17.7) | 2.35 (2.22, 2.49) | <0.0001 | 15,917 | 278 (1.7%) | 1.7 (1.6, 2) | 0.22 (0.19, 0.25) | <0.0001 |

Viral load measurements were not routinely collected in the 2012 survey round and we are therefore unable to estimate the prevalence of suppression among people living with HIV (PLHIV) for this time frame. PLHIV with missing viral load data in later survey rounds were dropped when estimating the prevalence of suppressed HIV. Estimates were generated using Poisson regression with robust standard errors with survey round as a predictor variable. Generalized estimating equations with correlation structure selection by Quasi Information Criterion value (PLHIV, viremic PLHIV: independence) were used to account for repeat participants across study rounds. 95% confidence intervals indicate the Wald confidence interval around the mean value in each category. *p*-values that coefficients are different from 0 at the  level were calculated using the Wald method.

**Supplementary Table 7: Prevalence of treatment-experience and suppression among RCCS participants living with HIV.**

| | | Treatment-experienced PLHIV | | | | Suppressed PLHIV | | | | |
|---|---|---|---|---|---|---|---|---|---|---|
| **Survey round** | ***n*** | **Obs (%)** | **Prev. (95% CI)** | **Prev. ratio (95% CI)** | ***p*-value** | **n** | **Obs (%)** | **Prev. (95% CI)** | ***Prev. ratio (95% CI)*** | ***p*-value** |
| 2012 | 3,498 | 1,240 (35.4%) | 35.4 (33.9, 37.1) | ref | ref | | | | | |
| 2014 | 3,396 | 2,120 (62.4%) | 62.4 (60.8, 64.1) | 1.76 (1.68, 1.84) | <0.0001 | 3,359 | 1,917 (57.1%) | 57.1 (55.4, 58.8) | ref | ref |
| 2015 | 3,616 | 2,826 (78.2%) | 78.2 (76.8, 79.5) | 2.2 (2.11, 2.31) | <0.0001 | 3,610 | 2,580 (71.5%) | 71.5 (70, 73) | 1.25 (1.21, 1.29) | <0.0001 |
| 2017 | 3,633 | 3,143 (86.5%) | 86.5 (85.4, 87.6) | 2.44 (2.33, 2.56) | <0.0001 | 3,623 | 2,897 (80%) | 80 (78.7, 81.3) | 1.4 (1.36, 1.45) | <0.0001 |
| 2019 | 3,347 | 3,064 (91.5%) | 91.5 (90.6, 92.5) | 2.58 (2.47, 2.7) | <0.0001 | 3,333 | 2,880 (86.4%) | 86.4 (85.3, 87.6) | 1.51 (1.47, 1.56) | <0.0001 |
| 2022 | 2,893 | 2,731 (94.4%) | 94.4 (93.6, 95.2) | 2.66 (2.54, 2.79) | <0.0001 | 2,863 | 2,585 (90.3%) | 90.3 (89.2, 91.4) | 1.58 (1.53, 1.63) | <0.0001 |

Viral load measurements were not routinely collected in the 2012 survey round and we are therefore unable to estimate the prevalence of suppression among people living with HIV (PLHIV) for this time frame. PLHIV with missing viral load data in later survey rounds were dropped when estimating the prevalence of suppressed HIV. Estimates were generated using Poisson regression with robust standard errors with survey round as a predictor variable. Generalized estimating equations with correlation structure selection by Quasi Information Criterion value (treatment-experienced PLHIV, suppressed PLHIV: independence) were used to account for repeat participants across study rounds. 95% confidence intervals indicate the Wald confidence interval around the mean value in each category. *p*-values that coefficients are different from 0 at the  level were calculated using the Wald method.

**Supplementary Table 8: Prevalence of suppression among treatment-experienced RCCS participants living with HIV**

| | | Suppressed HIV (≤1,000 copies/mL) | | | | 401-1000 copies/mL | | | |
|---|---|---|---|---|---|---|---|---|---|
| Survey round | *n* | Obs (%) | Prev. (95% CI) | Prev. ratio (95% CI) | *p*-value | Obs (%) | Prev. (95% CI) | Prev. ratio (95% CI) | *p*-value |
| 2014 | 2,110 | 1,917 (90.9%) | 91 (89.8, 92.1) | ref | ref | 102 (4.8%) | 5 (4.2, 6) | ref | ref |
| 2015 | 2,821 | 2,580 (91.5%) | 91.1 (90.1, 92.1) | 1 (0.99, 1.02) | 0.85 | 92 (3.3%) | 3.3 (2.7, 4.1) | 0.67 (0.52, 0.86) | 0.0017 |
| 2017 | 3,136 | 2,897 (92.4%) | 91.9 (91, 92.7) | 1.01 (0.99, 1.02) | 0.2 | 59 (1.9%) | 1.9 (1.5, 2.5) | 0.39 (0.29, 0.52) | <0.0001 |
| 2019 | 3,053 | 2,880 (94.3%) | 93.6 (92.8, 94.4) | 1.03 (1.01, 1.04) | 0.00015 | 46 (1.5%) | 1.6 (1.2, 2.1) | 0.31 (0.22, 0.43) | <0.0001 |
| 2022 | 2,702 | 2,585 (95.7%) | 95.4 (94.6, 96.1) | 1.05 (1.03, 1.06) | <0.0001 | 36 (1.3%) | 1.3 (1, 1.9) | 0.27 (0.19, 0.39) | <0.0001 |

| | | 201-400 copies/mL | | | | 0-200 copies/mL | | | |
|---|---|---|---|---|---|---|---|---|---|
| Survey round | *n* | Obs (%) | Prev. (95% CI) | Prev. ratio (95% CI) | *p*-value | Obs (%) | Prev. (95% CI) | Prev. ratio (95% CI) | *p*-value |
| 2014 | 2,110 | 62 (2.9%) | 3 (2.3, 3.8) | ref | ref | 1,753 (83.1%) | 83.1 (81.5, 84.7) | ref | ref |
| 2015 | 2,821 | 37 (1.3%) | 1.3 (1, 1.8) | 0.45 (0.31, 0.66) | <0.0001 | 2,451 (86.9%) | 86.9 (85.6, 88.1) | 1.05 (1.02, 1.07) | <0.0001 |
| 2017 | 3,136 | 38 (1.2%) | 1.2 (0.9, 1.7) | 0.41 (0.28, 0.61) | <0.0001 | 2,800 (89.3%) | 89.3 (88.2, 90.4) | 1.07 (1.05, 1.1) | <0.0001 |
| 2019 | 3,053 | 36 (1.2%) | 1.2 (0.9, 1.6) | 0.4 (0.27, 0.6) | <0.0001 | 2,798 (91.6%) | 91.6 (90.7, 92.6) | 1.1 (1.08, 1.13) | <0.0001 |
| 2022 | 2,702 | 21 (0.8%) | 0.8 (0.5, 1.2) | 0.26 (0.16, 0.43) | <0.0001 | 2,528 (93.6%) | 93.6 (92.6, 94.5) | 1.13 (1.1, 1.15) | <0.0001 |

Viral load measurements were not routinely collected in the 2012 survey round and we are therefore unable to estimate the prevalence of suppression among treatment-experienced people living with HIV (PLHIV) for this time frame. PLHIV with missing viral load data in later survey rounds were dropped when estimating the prevalence of suppressed HIV. Estimates were generated using Poisson regression with robust standard errors with survey round as a predictor variable. Generalized estimating equations with correlation structure selection by Quasi Information Criterion value (Suppressed: AR1, 401-1000 copies/mL: exchangeable, 201-400 copies/mL: exchangeable, 0-200 copies/mL: independence) were used to account for repeat participants across study rounds. 95% confidence intervals indicate the Wald confidence interval around the mean value in each category. *p*-values that coefficients are different from 0 at the  level were calculated using the Wald method.

**Supplementary Table 9: List of surveyed clinics serving PLHIV who participated in the 2022 Rakai Community Cohort Study survey round.**

| Clinic name | 2022 RCCS participants w/ HIV | |
|---|---|---|
| | *n* | % |
| Kasensero | 660 | 25.6 |
| Rakai Program Kalisizo Hub | 336 | 13.1 |
| Kakuuto | 159 | 6.2 |
| Kasasa | 120 | 4.7 |
| Rakai hospital | 101 | 3.9 |
| Taso Masaka | 93 | 3.6 |
| Kabira | 92 | 3.6 |
| Kasaali | 79 | 3.1 |
| Lwanda | 79 | 3.1 |
| Kifamba | 65 | 2.5 |
| Kamulegu H/C III | 39 | 1.5 |
| Mitukula | 35 | 1.4 |
| Lyantonde Hospital | 25 | 1 |
| Kyanamukaaka HC IV | 24 | 0.9 |

**Supplementary Table 10: Proportion of care seeking PLHIV prescribed DTG at surveyed clinics stratified by date and sex.**

| Quarter | Men | | | Women | | |
|---|---|---|---|---|---|---|
| | *n* | DTG | % (95% CI) | n | DTG | % (95% CI) |
| 2018 Q1 | 7,588 | 985 | 13 (12.2, 13.7) | 13,968 | 2,489 | 17.8 (17.2, 18.5) |
| 2018 Q2 | 7,879 | 1,142 | 14.5 (13.7, 15.3) | 14,465 | 2,735 | 18.9 (18.3, 19.5) |
| 2018 Q3 | 8,152 | 1,274 | 15.6 (14.8, 16.4) | 14,821 | 2,860 | 19.3 (18.7, 19.9) |
| 2018 Q4 | 8,105 | 2,051 | 25.3 (24.4, 26.3) | 14,584 | 3,571 | 24.5 (23.8, 25.2) |
| 2019 Q1 | 8,279 | 2,513 | 30.4 (29.4, 31.3) | 15,456 | 3,880 | 25.1 (24.4, 25.8) |
| 2019 Q2 | 9,398 | 3,323 | 35.4 (34.4, 36.3) | 15,626 | 4,428 | 28.3 (27.6, 29) |
| 2019 Q3 | 8,789 | 3,371 | 38.4 (37.3, 39.4) | 15,939 | 5,200 | 32.6 (31.9, 33.4) |
| 2019 Q4 | 8,773 | 5,683 | 64.8 (63.8, 65.8) | 15,962 | 7,565 | 47.4 (46.6, 48.2) |
| 2020 Q1 | 8,631 | 7,286 | 84.4 (83.7, 85.2) | 16,423 | 10,611 | 64.6 (63.9, 65.3) |
| 2020 Q2 | 8,702 | 7,434 | 85.4 (84.7, 86.2) | 16,173 | 10,501 | 64.9 (64.2, 65.7) |
| 2020 Q3 | 9,317 | 8,043 | 86.3 (85.6, 87) | 16,408 | 11,156 | 68 (67.3, 68.7) |
| 2020 Q4 | 9,724 | 8,746 | 89.9 (89.3, 90.5) | 17,012 | 13,046 | 76.7 (76.1, 77.3) |
| 2021 Q1 | 9,565 | 8,932 | 93.4 (92.9, 93.9) | 17,561 | 15,347 | 87.4 (86.9, 87.9) |
| 2021 Q2 | 10,361 | 9,886 | 95.4 (95, 95.8) | 17,100 | 15,689 | 91.7 (91.3, 92.2) |
| 2021 Q3 | 9,930 | 9,513 | 95.8 (95.4, 96.2) | 17,803 | 16,748 | 94.1 (93.7, 94.4) |
| 2021 Q4 | 9,507 | 9,202 | 96.8 (96.4, 97.1) | 17,135 | 16,390 | 95.7 (95.3, 96) |
| 2022 Q1 | 9,824 | 9,559 | 97.3 (97, 97.6) | 17,549 | 17,003 | 96.9 (96.6, 97.1) |
| 2022 Q2 | 9,667 | 9,431 | 97.6 (97.3, 97.9) | 17,813 | 17,080 | 95.9 (95.6, 96.2) |
| 2022 Q3 | 9,800 | 9,579 | 97.7 (97.5, 98) | 17,617 | 17,266 | 98 (97.8, 98.2) |
| 2022 Q4 | 9,658 | 9,488 | 98.2 (98, 98.5) | 17,591 | 17,283 | 98.2 (98.1, 98.4) |
| 2023 Q1 | 9,715 | 9,519 | 98 (97.7, 98.3) | 17,882 | 17,312 | 96.8 (96.6, 97.1) |

95% confidence intervals indicate the Wald confidence interval around the mean value in each category.

**Supplementary Table 11: Proportion of Treatment-experienced viremic participant visits on which deep-sequence based prediction of drug resistance was attempted and successful.**

| | | Treatment-experienced viremic PLHIV | | | | | | | |
|---|---|---|---|---|---|---|---|---|---|
| | | Participant-visits | Sequenced (% of par.-visits) | Deep-sequence based drug resistance prediction (% of sequenced) | | | | | |
| | | | | Any | INSTI | NNRTI | NRTI | PI | all |
| Overall | | 963 | 936 (97.2%) | 783 (83.7%) | 770 (82.3%) | 677 (72.3%) | 656 (70.1%) | 711 (76%) | 630 (67.3%) |
| Survey round | | | | | | | | | |
| | 2014 | 193 | 182 (94.3%) | 94 (51.6%) | 90 (49.5%) | 66 (36.3%) | 65 (35.7%) | 76 (41.8%) | 57 (31.3%) |
| | 2015 | 241 | 233 (96.7%) | 215 (92.3%) | 213 (91.4%) | 192 (82.4%) | 180 (77.3%) | 190 (81.5%) | 172 (73.8%) |
| | 2017 | 239 | 235 (98.3%) | 218 (92.8%) | 218 (92.8%) | 210 (89.4%) | 207 (88.1%) | 214 (91.1%) | 205 (87.2%) |
| | 2019 | 173 | 170 (98.3%) | 147 (86.5%) | 141 (82.9%) | 119 (70%) | 119 (70%) | 134 (78.8%) | 112 (65.9%) |
| | 2022 | 117 | 116 (99.1%) | 109 (94%) | 108 (93.1%) | 90 (77.6%) | 85 (73.3%) | 97 (83.6%) | 84 (72.4%) |
| Sequencing approach | | | | | | | | | |
| | amplicon | 9 | 9 (100%) | 6 (66.7%) | 5 (55.6%) | 5 (55.6%) | 6 (66.7%) | 6 (66.7%) | 4 (44.4%) |
| | veSeq-HIV | 927 | 927 (100%) | 777 (83.8%) | 765 (82.5%) | 672 (72.5%) | 650 (70.1%) | 705 (76.1%) | 626 (67.5%) |
| Subtype | | 32 | 5 (15.6%) | | | | | | |
| | A1 | 255 | 255 (100%) | 218 (85.5%) | 215 (84.3%) | 182 (71.4%) | 182 (71.4%) | 195 (76.5%) | 175 (68.6%) |
| | C | 32 | 32 (100%) | 28 (87.5%) | 28 (87.5%) | 23 (71.9%) | 23 (71.9%) | 23 (71.9%) | 21 (65.6%) |
| | D | 505 | 505 (100%) | 426 (84.4%) | 421 (83.4%) | 375 (74.3%) | 363 (71.9%) | 395 (78.2%) | 348 (68.9%) |
| | other | 139 | 139 (100%) | 111 (79.9%) | 106 (76.3%) | 97 (69.8%) | 88 (63.3%) | 98 (70.5%) | 86 (61.9%) |
| | missing | 32 | 5 (15.6%) | NA (NA%) | NA (NA%) | NA (NA%) | NA (NA%) | NA (NA%) | NA (NA%) |
| Viral load (log10 copies/mL) | | | | | | | | | |
| | (3,4] | 500 | 484 (96.8%) | 359 (74.2%) | 348 (71.9%) | 280 (57.9%) | 257 (53.1%) | 296 (61.2%) | 241 (49.8%) |
| | (4,5] | 364 | 357 (98.1%) | 330 (92.4%) | 328 (91.9%) | 304 (85.2%) | 305 (85.4%) | 321 (89.9%) | 296 (82.9%) |
| | (5, Inf] | 99 | 95 (96%) | 94 (98.9%) | 94 (98.9%) | 93 (97.9%) | 94 (98.9%) | 94 (98.9%) | 93 (97.9%) |

**Supplementary Table 12: Prevalence of resistance among treatment-experienced viremic PLHIV in the Rakai Community Cohort Study, 2014 - 2022.**

| | Any | | | | | INSTI | | | | | NNRTI | | | | |
|---|---|---|---|---|---|---|---|---|---|---|---|---|---|---|---|
| **Survey round** | ***n*** | **Obs (%)** | **Prev. (95% CI)** | **Prev. ratio (95% CI)** | ***p*-value** | ***n*** | **Obs (%)** | **Prev. (95% CI)** | **Prev. ratio (95% CI)** | ***p*-value** | ***n*** | **Obs (%)** | **Prev. (95% CI)** | **Prev. ratio (95% CI)** | ***p*-value** |
| 2014 | 62 | 29 (46.8%) | 51.1 (40.7, 64.2) | ref | ref | 90 | 0 (0%) | 0 (NA, NA) | ref | ref | 66 | 28 (42.4%) | 46 (35.9, 58.9) | ref | ref |
| 2015 | 180 | 95 (52.8%) | 49.8 (44.2, 56.1) | 0.98 (0.78, 1.22) | 0.83 | 213 | 1 (0.5%) | 0.4 (0.1, 3) | NA (0, Inf) | | 192 | 92 (47.9%) | 46 (40.7, 52.1) | 1 (0.79, 1.27) | 0.99 |
| 2017 | 207 | 95 (45.9%) | 44.4 (39.1, 50.4) | 0.87 (0.68, 1.11) | 0.26 | 218 | 2 (0.9%) | 0.9 (0.2, 3.5) | NA (0, Inf) | | 210 | 91 (43.3%) | 43.4 (38.3, 49.1) | 0.94 (0.73, 1.22) | 0.66 |
| 2019 | 114 | 50 (43.9%) | 41.4 (34.2, 50.1) | 0.81 (0.6, 1.09) | 0.16 | 141 | 0 (0%) | 0 (0, 0) | NA (0, Inf) | | 119 | 47 (39.5%) | 37.6 (31.3, 45.3) | 0.82 (0.61, 1.11) | 0.19 |
| 2022 | 85 | 17 (20%) | 27.9 (21.3, 36.5) | 0.55 (0.39, 0.77) | 0.00066 | 108 | 3 (2.8%) | 2.7 (0.9, 8.3) | NA (0, Inf) | | 90 | 14 (15.6%) | 22.2 (17.1, 28.9) | 0.48 (0.34, 0.69) | <0.0001 |

| | NRTI | | | | | PI | | | | |
|---|---|---|---|---|---|---|---|---|---|---|
| **Survey round** | ***n*** | **Obs (%)** | **Prev. (95% CI)** | **Prev. ratio (95% CI)** | ***p*-value** | ***n*** | **Obs (%)** | **Prev. (95% CI)** | **Prev. ratio (95% CI)** | ***p*-value** |
| 2014 | 65 | 25 (38.5%) | 34.1 (25.4, 45.7) | ref | ref | 76 | 0 (0%) | 0 (0, Inf) | ref | ref |
| 2015 | 180 | 77 (42.8%) | 38.9 (33.4, 45.4) | 1.14 (0.86, 1.52) | 0.35 | 190 | 6 (3.2%) | 3.1 (1.4, 6.6) | NA (0, Inf) | 1 |
| 2017 | 207 | 72 (34.8%) | 33 (28, 38.8) | 0.97 (0.71, 1.33) | 0.84 | 214 | 5 (2.3%) | 2.2 (1, 5.3) | NA (0, Inf) | 1 |
| 2019 | 119 | 29 (24.4%) | 24.5 (18.3, 32.8) | 0.72 (0.48, 1.08) | 0.11 | 134 | 5 (3.7%) | 3.8 (1.6, 9) | NA (0, Inf) | 1 |
| 2022 | 85 | 6 (7.1%) | 12.3 (7.7, 19.9) | 0.36 (0.21, 0.63) | 0.00035 | 97 | 1 (1%) | 0.9 (0.1, 6) | NA (0, Inf) | 1 |

Viral load measurements were not routinely collected in the 2012 survey round and therefore we exclude this this time frame. Estimates were generated using Poisson regression with robust standard errors with survey round as a predictor variable. Generalized estimating equations with correlation structure selection by Quasi Information Criterion value (Any, NRTI, NNRTI: AR1) were used to account for repeat participants across study rounds. 95% confidence intervals indicate the Wald confidence interval around the mean value in each category. *p*-values that coefficients are different from 0 at the  level were calculated using the Wald method. INSTI = integrase strand transfer inhibitor, NNRTI = non-nucleoside reverse transcriptase inhibitor, NRTI = nucleoside reverse transcriptase inhibitor, PI = protease inhibitor.

**Supplementary Table 13: Demographics of treatment-experienced viremic people living with HIV with resistance to any drug classes in the Rakai Community Cohort Study, 2014-2022.**

| Variable | Viremic treatment-experienced PLHIV | | Risk ratio (95% CI) | *p*-value | Adj. risk ratio | Adj. *p*-value |
|---|---|---|---|---|---|---|
| | Genotyped | W/ resistance (%) | | | | |
| **Age** | | | | | | |
| (14,24] | 98 | 50 (51%) | ref | ref | ref | ref |
| (24,34] | 296 | 128 (43.2%) | 0.81 (0.62, 1.06) | 0.12 | 0.83 (0.64, 1.08) | 0.17 |
| (34,49] | 254 | 108 (42.5%) | 0.8 (0.6, 1.06) | 0.13 | 0.81 (0.61, 1.08) | 0.15 |
| **Community type** | | | | | | |
| fishing | 353 | 147 (41.6%) | ref | ref | ref | ref |
| inland | 295 | 139 (47.1%) | 1.22 (0.98, 1.51) | 0.07 | 1.23 (1, 1.51) | 0.053 |
| **Sex** | | | | | | |
| female | 346 | 177 (51.2%) | ref | ref | ref | ref |
| male | 302 | 109 (36.1%) | 0.65 (0.52, 0.82) | 0.0002 | 0.65 (0.53, 0.81) | 0.0001 |
| **Subtype** | | | | | | |
| A1 | 180 | 81 (45%) | ref | ref | ref | ref |
| C | 23 | 6 (26.1%) | 0.67 (0.36, 1.25) | 0.2 | 0.67 (0.36, 1.24) | 0.2 |
| D | 358 | 160 (44.7%) | 0.9 (0.71, 1.12) | 0.34 | 0.88 (0.7, 1.1) | 0.26 |
| other | 87 | 39 (44.8%) | 1.11 (0.82, 1.5) | 0.49 | 1.06 (0.8, 1.41) | 0.67 |

*P*-values are calculated from either univariate or bivariate (with survey round) Poisson regression with robust standard errors. Generalized estimating equations with correlation structure selection by Quasi Information Criterion value (age, community type, sex, subtype: AR1) were used to account for repeat participants across study rounds.

**Supplementary Table 14: Prevalence of multi-class resistance among treatment-experienced viremic PLHIV in the Rakai Community Cohort Study, 2014 - 2022.**

| | Multi-class | | | | | Single-class | | | | |
|---|---|---|---|---|---|---|---|---|---|---|
| **Survey round** | ***n*** | **Obs (%)** | **Prev. (95% CI)** | **Prev. ratio (95% CI)** | ***p*-value** | ***n*** | **Obs (%)** | **Prev. (95% CI)** | **Prev. ratio (95% CI)** | ***p*-value** |
| 2014 | 57 | 23 (40.4%) | 41.5 (31.2, 55.3) | ref | ref | 57 | 1 (1.8%) | 2.3 (0.4, 14.3) | ref | ref |
| 2015 | 172 | 72 (41.9%) | 38.4 (33, 44.8) | 0.92 (0.7, 1.22) | 0.58 | 172 | 15 (8.7%) | 8.5 (5.3, 13.4) | 3.69 (0.57, 23.71) | 0.17 |
| 2017 | 205 | 72 (35.1%) | 33.5 (28.4, 39.4) | 0.81 (0.59, 1.1) | 0.17 | 205 | 21 (10.2%) | 10.3 (7, 15.3) | 4.51 (0.7, 29.26) | 0.11 |
| 2019 | 112 | 28 (25%) | 25.5 (19.7, 33.1) | 0.61 (0.42, 0.9) | 0.013 | 112 | 20 (17.9%) | 17.9 (12.1, 26.3) | 7.8 (1.2, 50.68) | 0.031 |
| 2022 | 84 | 5 (6%) | 9.7 (6.1, 15.5) | 0.23 (0.14, 0.4) | <0.0001 | 84 | 11 (13.1%) | 12.9 (7.5, 21.9) | 5.62 (0.83, 37.8) | 0.076 |

Viral load measurements were not routinely collected in the 2012 survey round and therefore we exclude this this time frame. Estimates were generated using Poisson regression with robust standard errors with survey round as a predictor variable. Multi-class refers to any combination of simultaneous resistance to any of the four drug classes. Generalized estimating equations with correlation structure selection by Quasi Information Criterion value (multi-class: AR1, single-class: exchangeable) were used to account for repeat participants across study rounds. 95% confidence intervals indicate the Wald confidence interval around the mean value in each category. *p*-values that coefficients are different from 0 at the  level were calculated using the Wald method.

**Supplementary Table 15: Prevalence of resistance-conferring mutations among treatment-experienced viremic PLHIV in the Rakai Community Cohort Study, 2014 - 2022.**

| | | inT97A | | | | rtK103N | | | | inS153Y | | | |
|---|---|---|---|---|---|---|---|---|---|---|---|---|---|
| Survey round | *n* | Obs (%) | Prev. (95% CI) | Prev. ratio (95% CI) | *p*-value | Obs (%) | Prev. (95% CI) | Prev. ratio (95% CI) | *p*-value | Obs (%) | Prev. (95% CI) | Prev. ratio (95% CI) | *p*-value |
| 2014 | 134 | 2 (1.5%) | 8.2 (2.2, 30.6) | ref | ref | 10 (7.5%) | 25 (14, 44.7) | ref | ref | 0 (0%) | 0 (0, Inf) | ref | ref |
| 2015 | 192 | 15 (7.8%) | 9.7 (5.9, 15.7) | 1.18 (0.29, 4.76) | 0.82 | 46 (24%) | 32.8 (25.7, 41.8) | 1.31 (0.71, 2.42) | 0.39 | 0 (0%) | 0 (0, 0) | NA (0, Inf) | 1 |
| 2017 | 212 | 21 (9.9%) | 11.6 (7.7, 17.4) | 1.41 (0.36, 5.57) | 0.62 | 50 (23.6%) | 27 (21.3, 34.3) | 1.08 (0.58, 2.01) | 0.8 | 0 (0%) | 0 (NA, NA) | NA (0, Inf) | 1 |
| 2019 | 159 | 5 (3.1%) | 4.5 (1.9, 10.7) | 0.55 (0.11, 2.64) | 0.45 | 29 (18.2%) | 26.3 (19.1, 36.2) | 1.05 (0.54, 2.04) | 0.88 | 0 (0%) | 0 (0, 0) | NA (0, Inf) | 1 |
| 2022 | 108 | 9 (8.3%) | 10.2 (5.4, 19.1) | 1.24 (0.29, 5.31) | 0.77 | 8 (7.4%) | 9.1 (4.6, 18) | 0.37 (0.15, 0.89) | 0.027 | 7 (6.5%) | 7.8 (3.9, 15.7) | NA (0, Inf) | 1 |

| | | prL23I | | | | rtM184V | | | | rtM41L | | | |
|---|---|---|---|---|---|---|---|---|---|---|---|---|---|
| Survey round | *n* | Obs (%) | Prev. (95% CI) | Prev. ratio (95% CI) | *p*-value | Obs (%) | Prev. (95% CI) | Prev. ratio (95% CI) | *p*-value | Obs (%) | Prev. (95% CI) | Prev. ratio (95% CI) | *p*-value |
| 2014 | 134 | 0 (0%) | 0 (NA, NA) | ref | ref | 19 (14.2%) | 47.3 (33.7, 66.4) | ref | ref | 5 (3.7%) | 11.1 (4.6, 26.7) | ref | ref |
| 2015 | 192 | 0 (0%) | 0 (0, 0) | NA (0, Inf) | <0.0001 | 59 (30.7%) | 42.1 (34.5, 51.4) | 0.89 (0.61, 1.3) | 0.55 | 11 (5.7%) | 7.8 (4.4, 14.1) | 0.71 (0.27, 1.85) | 0.48 |
| 2017 | 212 | 0 (0%) | 0 (0, 0) | NA (0, Inf) | <0.0001 | 65 (30.7%) | 35 (28.7, 42.7) | 0.74 (0.5, 1.09) | 0.13 | 11 (5.2%) | 5.8 (3.3, 10.4) | 0.53 (0.18, 1.51) | 0.23 |
| 2019 | 159 | 0 (0%) | 0 (0, 0) | NA (0, Inf) | <0.0001 | 24 (15.1%) | 22 (15.3, 31.6) | 0.47 (0.28, 0.77) | 0.0026 | 4 (2.5%) | 3.7 (1.4, 10) | 0.33 (0.09, 1.26) | 0.11 |
| 2022 | 108 | 5 (4.6%) | 6.6 (2.8, 15.2) | NA (0, Inf) | <0.0001 | 4 (3.7%) | 5.1 (2, 13.3) | 0.11 (0.04, 0.3) | <0.0001 | 4 (3.7%) | 5 (1.9, 13) | 0.45 (0.12, 1.66) | 0.23 |

| | | rtP225H | | | | rtT215Y | | | | rtY181C | | | |
|---|---|---|---|---|---|---|---|---|---|---|---|---|---|
| Survey round | *n* | Obs (%) | Prev. (95% CI) | Prev. ratio (95% CI) | *p*-value | Obs (%) | Prev. (95% CI) | Prev. ratio (95% CI) | *p*-value | Obs (%) | Prev. (95% CI) | Prev. ratio (95% CI) | *p*-value |
| 2014 | 134 | 1 (0.75%) | 1.92 (0.27, 13.53) | ref | ref | 4 (2.99%) | 11.59 (4.31, 31.19) | ref | ref | 12 (8.96%) | 26.59 (15.68, 45.1) | ref | ref |
| 2015 | 192 | 6 (3.12%) | 4.05 (1.84, 8.93) | 2.11 (0.26, 17.23) | 0.49 | 5 (2.6%) | 4.28 (1.78, 10.28) | 0.37 (0.1, 1.38) | 0.14 | 33 (17.19 %) | 24.34 (17.96, 33) | 0.92 (0.51, 1.63) | 0.76 |

| | | | | | | | | | | | | | |
|---|---|---|---|---|---|---|---|---|---|---|---|---|---|
| 2017 | 212 | 11 (5.19%) | 6.14 (3.44, 10.95) | 3.2 (0.42, 24.43) | 0.26 | 5 (2.36%) | 2.76 (1.16, 6.58) | 0.24 (0.06, 0.89) | 0.032 | 27 (12.74 %) | 14.22 (10, 20.23) | 0.53 (0.29, 1) | 0.049 |
| 2019 | 159 | 9 (5.66%) | 8.48 (4.5, 16) | 4.42 (0.57, 34.35) | 0.16 | 2 (1.26%) | 2.04 (0.5, 8.37) | 0.18 (0.03, 0.98) | 0.048 | 10 (6.29%) | 9.72 (5.34, 17.66) | 0.37 (0.16, 0.81) | 0.014 |
| 2022 | 108 | 3 (2.78%) | 3.55 (1.15, 10.98) | 1.85 (0.19, 17.55) | 0.59 | 3 (2.78%) | 3.89 (1.28, 11.83) | 0.34 (0.08, 1.48) | 0.15 | 3 (2.78%) | 3.63 (1.18, 11.13) | 0.14 (0.04, 0.47) | 0.0016 |

| | | **inE138K** | | | |
|---|---|---|---|---|---|
| **Survey round** | ***n*** | **Obs (%)** | **Prev. (95% CI)** | **Prev. ratio (95% CI)** | ***p-value*** |
| 2014 | 134 | 0 (0%) | 0 (0, Inf) | ref | ref |
| 2015 | 192 | 1 (0.52%) | 0.62 (0.09, 4.41) | NA (0, Inf) | 1 |
| 2017 | 212 | 0 (0%) | 0 (0, 0) | NA (0, Inf) | 1 |
| 2019 | 159 | 0 (0%) | 0 (0, 0) | NA (0, Inf) | 1 |
| 2022 | 108 | 1 (0.93%) | 1.23 (0.18, 8.66) | NA (0, Inf) | 1 |

Viral load measurements were not routinely collected in the 2012 survey round and therefore we exclude this this time frame. Estimates were generated using Poisson regression with robust standard errors with survey round as a predictor variable. Multi-class refers to any combination of simultaneous resistance to any of the four drug classes. Generalized estimating equations with independent correlation structure to aid convergence for rare mutations were used. 95% confidence intervals indicate the Wald confidence interval around the mean value in each category. *p*-values that coefficients are different from 0 at the  level were calculated using the Wald method.

**Supplementary Table 16: Proportion of pre-treatment viremic participant visits on which deep-sequence based prediction of drug resistance was attempted and successful.**

| | | Pre-treatment viremic PLHIV | | | | | | | |
|---|---|---|---|---|---|---|---|---|---|
| | | Participant-visits | Sequenced (% of par.-visits) | Deep-sequence based drug resistance prediction (% of sequenced) | | | | | |
| | | | | Any | INSTI | NNRTI | NRTI | PI | all |
| Overall | | 4,981 | 4,924 (98.9%) | 3,918 (79.6%) | 2,766 (56.2%) | 3,206 (65.1%) | 3,176 (64.5%) | 3,744 (76%) | 2,461 (50%) |
| Survey round | | | | | | | | | |
| | 2012 | 2,015 | 1,993 (98.9%) | 1,420 (71.2%) | 516 (25.9%) | 979 (49.1%) | 1,017 (51%) | 1,387 (69.6%) | 482 (24.2%) |
| | 2014 | 1,249 | 1,228 (98.3%) | 856 (69.7%) | 636 (51.8%) | 668 (54.4%) | 659 (53.7%) | 774 (63%) | 502 (40.9%) |
| | 2015 | 789 | 784 (99.4%) | 757 (96.6%) | 737 (94%) | 719 (91.7%) | 687 (87.6%) | 722 (92.1%) | 667 (85.1%) |
| | 2017 | 487 | 481 (98.8%) | 470 (97.7%) | 465 (96.7%) | 459 (95.4%) | 450 (93.6%) | 465 (96.7%) | 450 (93.6%) |
| | 2019 | 280 | 279 (99.6%) | 264 (94.6%) | 262 (93.9%) | 238 (85.3%) | 227 (81.4%) | 253 (90.7%) | 227 (81.4%) |
| | 2022 | 161 | 159 (98.8%) | 151 (95%) | 150 (94.3%) | 143 (89.9%) | 136 (85.5%) | 143 (89.9%) | 133 (83.6%) |
| Sequencing approach | | | | | | | | | |
| | amplicon | 2,571 | 2,571 (100%) | 1,792 (69.7%) | 666 (25.9%) | 1,276 (49.6%) | 1,332 (51.8%) | 1,759 (68.4%) | 638 (24.8%) |
| | veSeq-HIV | 2,353 | 2,353 (100%) | 2,126 (90.4%) | 2,100 (89.2%) | 1,930 (82%) | 1,844 (78.4%) | 1,985 (84.4%) | 1,823 (77.5%) |
| Subtype | | | | | | | | | |
| | A1 | 1,356 | 1,356 (100%) | 1,200 (88.5%) | 851 (62.8%) | 1,010 (74.5%) | 1,004 (74%) | 1,140 (84.1%) | 758 (55.9%) |
| | C | 149 | 149 (100%) | 122 (81.9%) | 93 (62.4%) | 103 (69.1%) | 104 (69.8%) | 117 (78.5%) | 85 (57%) |
| | D | 1,908 | 1,908 (100%) | 1,623 (85.1%) | 1,338 (70.1%) | 1,439 (75.4%) | 1,400 (73.4%) | 1,550 (81.2%) | 1,189 (62.3%) |
| | other | 1,504 | 1,504 (100%) | 973 (64.7%) | 484 (32.2%) | 654 (43.5%) | 668 (44.4%) | 937 (62.3%) | 429 (28.5%) |
| | missing | 64 | 7 (10.9%) | | | | | | |
| Viral load (log10 copies/mL) | | | | | | | | | |
| | (3,4] | 1,677 | 1,653 (98.6%) | 1,076 (65.1%) | 711 (43%) | 774 (46.8%) | 739 (44.7%) | 980 (59.3%) | 551 (33.3%) |
| | (4,5] | 1,971 | 1,952 (99%) | 1,753 (89.8%) | 1,351 (69.2%) | 1,513 (77.5%) | 1,505 (77.1%) | 1,696 (86.9%) | 1,227 (62.9%) |
| | (5, Inf] | 541 | 537 (99.3%) | 525 (97.8%) | 469 (87.3%) | 502 (93.5%) | 503 (93.7%) | 520 (96.8%) | 460 (85.7%) |
| | missing | 792 | 782 (98.7%) | 564 (72.1%) | 235 (30.1%) | 417 (53.3%) | 429 (54.9%) | 548 (70.1%) | 223 (28.5%) |

**Supplementary Table 17: Prevalence of resistance among pre-treatment PLHIV in the Rakai Community Cohort Study, 2012 - 2022.**

| | INSTI | | | | | NNRTI | | | | |
|---|---|---|---|---|---|---|---|---|---|---|
| **Survey round** | ***n*** | **Obs (%)** | **Prev. (95% CI)** | **Prev. ratio (95% CI)** | ***p*-value** | ***n*** | **Obs (%)** | **Prev. (95% CI)** | **Prev. ratio (95% CI)** | ***p*-value** |
| 2012 | 516 | 5 (1%) | 1 (0.4, 2.4) | ref | ref | 979 | 52 (5.3%) | 5.4 (4.3, 6.9) | ref | ref |
| 2014 | 636 | 12 (1.9%) | 2.1 (1.2, 3.8) | 2.17 (0.74, 6.33) | 0.16 | 668 | 40 (6%) | 6.3 (4.9, 8.1) | 1.16 (0.87, 1.56) | 0.31 |
| 2015 | 737 | 1 (0.1%) | 0.2 (0, 1.1) | 0.16 (0.02, 1.35) | 0.091 | 719 | 52 (7.2%) | 7.2 (5.7, 9.1) | 1.32 (0.96, 1.81) | 0.087 |
| 2017 | 465 | 0 (0%) | 0 (0, 0) | 0 (0, 0) | <0.0001 | 459 | 45 (9.8%) | 9.9 (7.8, 12.4) | 1.81 (1.3, 2.51) | 0.0004 |
| 2019 | 262 | 0 (0%) | 0 (0, 0) | 0 (0, 0) | <0.0001 | 238 | 27 (11.3%) | 11.5 (8.6, 15.5) | 2.12 (1.45, 3.09) | 0.0001 |
| 2022 | 150 | 0 (0%) | 0 (0, 0) | 0 (0, 0) | <0.0001 | 143 | 23 (16.1%) | 14.8 (9.7, 22.7) | 2.72 (1.67, 4.44) | <0.0001 |

| | NRTI | | | | | PI | | | | |
|---|---|---|---|---|---|---|---|---|---|---|
| **Survey round** | ***n*** | **Obs (%)** | **Prev. (95% CI)** | **Prev. ratio (95% CI)** | ***p*-value** | ***n*** | **Obs (%)** | **Prev. (95% CI)** | **Prev. ratio (95% CI)** | ***p*-value** |
| 2012 | 1,017 | 22 (2.2%) | 1.9 (1.2, 3) | ref | ref | 1,387 | 29 (2.1%) | 2.2 (1.5, 3.1) | ref | ref |
| 2014 | 659 | 10 (1.5%) | 1.7 (0.9, 3.2) | 0.93 (0.44, 1.97) | 0.84 | 774 | 9 (1.2%) | 1.3 (0.7, 2.4) | 0.6 (0.31, 1.17) | 0.14 |
| 2015 | 687 | 13 (1.9%) | 1.9 (1.1, 3.3) | 1.03 (0.49, 2.16) | 0.94 | 722 | 10 (1.4%) | 1.4 (0.8, 2.5) | 0.66 (0.35, 1.24) | 0.19 |
| 2017 | 450 | 7 (1.6%) | 1.7 (0.9, 3.2) | 0.91 (0.41, 2.04) | 0.82 | 465 | 8 (1.7%) | 1.7 (0.8, 3.2) | 0.76 (0.37, 1.58) | 0.47 |
| 2019 | 227 | 4 (1.8%) | 2 (1, 4) | 1.05 (0.45, 2.45) | 0.92 | 253 | 2 (0.8%) | 0.8 (0.2, 3.4) | 0.37 (0.08, 1.65) | 0.19 |
| 2022 | 136 | 1 (0.7%) | 0.9 (0.2, 4.3) | 0.5 (0.1, 2.48) | 0.4 | 143 | 1 (0.7%) | 0.8 (0.1, 5.2) | 0.38 (0.06, 2.47) | 0.31 |

Estimates were generated using Poisson regression with robust standard errors with survey round as a predictor variable. Generalized estimating equations with correlation structure selection by Quasi Information Criterion value (NRTI, PI: AR1, NRTI: exchangeable) were used to account for repeat participants across study rounds. 95% confidence intervals indicate the Wald confidence interval around the mean value in each category. *p*-values that coefficients are different from 0 at the  level were calculated using the Wald method. INSTI = integrase strand transfer inhibitor, NNRTI = non-nucleoside reverse transcriptase inhibitor, NRTI = nucleoside reverse transcriptase inhibitor, PI = protease inhibitor.

**Supplementary Table 18: Demographics of pre-treatment people living with HIV with resistance to NNRTIs in the Rakai Community Cohort Study, 2014-2022.**

| Variable | Viremic treatment-experienced PLHIV | | Risk ratio (95% CI) | *p*-value | Adj. risk ratio | Adj. *p*-value |
|---|---|---|---|---|---|---|
| | Genotyped | W/ resistance (%) | | | | |
| **Age** | | | | | | |
| (14,24] | 649 | 53 (8.2%) | ref | ref | ref | ref |
| (24,34] | 1603 | 117 (7.3%) | 0.9 (0.66, 1.22) | 0.5 | 0.92 (0.67, 1.24) | 0.58 |
| (34,49] | 954 | 69 (7.2%) | 0.94 (0.65, 1.35) | 0.72 | 0.94 (0.65, 1.35) | 0.74 |
| **Community type** | | | | | | |
| fishing | 1529 | 110 (7.2%) | ref | ref | ref | ref |
| inland | 1677 | 129 (7.7%) | 1.13 (0.84, 1.5) | 0.42 | 1.17 (0.88, 1.56) | 0.27 |
| **Sex** | | | | | | |
| female | 1566 | 135 (8.6%) | ref | ref | ref | ref |
| male | 1640 | 104 (6.3%) | 0.74 (0.56, 0.98) | 0.035 | 0.72 (0.55, 0.95) | 0.021 |
| **Subtype** | | | | | | |
| A1 | 1010 | 75 (7.4%) | ref | ref | ref | ref |
| C | 103 | 8 (7.8%) | 1 (0.47, 2.1) | 0.99 | 1.01 (0.39, 2.59) | 0.98 |
| D | 1439 | 114 (7.9%) | 1.13 (0.81, 1.58) | 0.46 | 1.12 (0.82, 1.54) | 0.47 |
| other | 654 | 42 (6.4%) | 0.9 (0.6, 1.35) | 0.6 | 1.04 (0.72, 1.48) | 0.85 |

*P*-values are calculated from either univariate or bivariate (with survey round) Poisson regression with robust standard errors. Generalized estimating equations with correlation structure selection by Quasi Information Criterion value (age, community type, sex, subtype: AR1) were used to account for repeat participants across study rounds. NNRTI = non-nucleoside reverse transcriptase inhibitor.

**Supplementary Table 19: Prevalence of resistance-conferring mutations among pre-treatment viremic PLHIV in the Rakai Community Cohort Study, 2012 - 2022.**

| | | rtK103N | | | | inT97A | | | | inS153Y | | | |
|---|---|---|---|---|---|---|---|---|---|---|---|---|---|
| **Survey round** | ***n*** | **Obs (%)** | **Prev. (95% CI)** | **Prev. ratio (95% CI)** | ***p*-value** | **Obs (%)** | **Prev. (95% CI)** | **Prev. ratio (95% CI)** | ***p*-value** | **Obs (%)** | **Prev. (95% CI)** | **Prev. ratio (95% CI)** | ***p*-value** |
| 2012 | 1,223 | 5 (0.4%) | 1.9 (0.8, 4.5) | ref | ref | 17 (1.4%) | 6.4 (4, 10.2) | ref | ref | 0 (0%) | 0 (NA, NA) | ref | ref |
| 2014 | 1,308 | 15 (1.1%) | 2.9 (1.7, 4.8) | 1.56 (0.59, 4.13) | 0.37 | 49 (3.7%) | 9.6 (7.3, 12.6) | 1.49 (0.89, 2.48) | 0.13 | 0 (0%) | 0 (0, 0) | NA (0, Inf) | <0.0001 |
| 2015 | 838 | 27 (3.2%) | 3.9 (2.7, 5.7) | 2.1 (0.81, 5.42) | 0.13 | 65 (7.8%) | 9.3 (7.4, 11.7) | 1.45 (0.87, 2.43) | 0.16 | 0 (0%) | 0 (0, 0) | NA (0, Inf) | <0.0001 |
| 2017 | 514 | 33 (6.4%) | 7 (5, 9.7) | 3.74 (1.47, 9.52) | 0.0056 | 48 (9.3%) | 10.2 (7.8, 13.3) | 1.59 (0.93, 2.72) | 0.091 | 0 (0%) | 0 (0, 0) | NA (0, Inf) | <0.0001 |
| 2019 | 294 | 17 (5.8%) | 7.5 (4.7, 12) | 4.04 (1.5, 10.85) | 0.0056 | 33 (11.2%) | 13.7 (10, 18.9) | 2.14 (1.22, 3.76) | 0.0083 | 0 (0%) | 0 (0, 0) | NA (0, Inf) | <0.0001 |
| 2022 | 170 | 14 (8.2%) | 10.2 (6.2, 17) | 5.49 (2, 15.06) | 0.00095 | 10 (5.9%) | 7 (3.8, 12.8) | 1.09 (0.51, 2.34) | 0.83 | 10 (5.9%) | 7.8 (4.3, 14.3) | NA (0, Inf) | <0.0001 |

| | | rtE138A | | | | prL23I | | | | prL33F | | | |
|---|---|---|---|---|---|---|---|---|---|---|---|---|---|
| **Survey round** | ***n*** | **Obs (%)** | **Prev. (95% CI)** | **Prev. ratio (95% CI)** | ***p*-value** | **Obs (%)** | **Prev. (95% CI)** | **Prev. ratio (95% CI)** | ***p*-value** | **Obs (%)** | **Prev. (95% CI)** | **Prev. ratio (95% CI)** | ***p*-value** |
| 2012 | 1,223 | 5 (0.4%) | 1.9 (0.8, 4.5) | ref | ref | 0 (0%) | 0 (0, Inf) | ref | ref | 3 (0.2%) | 1.2 (0.4, 3.6) | ref | ref |
| 2014 | 1,308 | 18 (1.4%) | 3.3 (2.1, 5.3) | 1.75 (0.73, 4.21) | 0.21 | 1 (0.1%) | 0.2 (0, 1.3) | NA (0, Inf) | 1 | 7 (0.5%) | 1.4 (0.7, 3.1) | 1.24 (0.37, 4.17) | 0.73 |
| 2015 | 838 | 26 (3.1%) | 3.7 (2.5, 5.4) | 1.97 (0.79, 4.94) | 0.15 | 0 (0%) | 0 (0, 0) | NA (0, Inf) | 1 | 7 (0.8%) | 1 (0.5, 2.1) | 0.86 (0.22, 3.32) | 0.82 |
| 2017 | 514 | 12 (2.3%) | 2.5 (1.4, 4.4) | 1.35 (0.48, 3.81) | 0.57 | 0 (0%) | 0 (0, 0) | NA (0, Inf) | 1 | 8 (1.6%) | 1.7 (0.9, 3.4) | 1.49 (0.4, 5.64) | 0.55 |
| 2019 | 294 | 10 (3.4%) | 4.2 (2.3, 7.7) | 2.24 (0.77, 6.52) | 0.14 | 0 (0%) | 0 (0, 0) | NA (0, Inf) | 1 | 5 (1.7%) | 2 (0.9, 4.9) | 1.76 (0.42, 7.37) | 0.44 |
| 2022 | 170 | 7 (4.1%) | 5.4 (2.6, 11.1) | 2.86 (0.92, 8.92) | 0.07 | 6 (3.5%) | 4.9 (2.2, 10.6) | NA (0, Inf) | 1 | 5 (2.9%) | 3.5 (1.5, 8.3) | 3.04 (0.73, 12.68) | 0.13 |

| | | prG73V | | | | rtF77L | | | | rtG190A | | | |
|---|---|---|---|---|---|---|---|---|---|---|---|---|---|
| **Survey round** | ***n*** | **Obs (%)** | **Prev. (95% CI)** | **Prev. ratio (95% CI)** | ***p*-value** | **Obs (%)** | **Prev. (95% CI)** | **Prev. ratio (95% CI)** | ***p*-value** | **Obs (%)** | **Prev. (95% CI)** | **Prev. ratio (95% CI)** | ***p*-value** |

| | | | | | | | | | | | | | |
|---|---|---|---|---|---|---|---|---|---|---|---|---|---|
| 2012 | 1223 | 0 (0%) | 0 (NA, NA) | ref | ref | 0 (0%) | 0 (NA, NA) | ref | ref | 2 (0.16%) | 0.87 (0.22, 3.44) | ref | ref |
| 2014 | 1308 | 0 (0%) | 0 (0, 0) | NA (0, Inf) | <0.0001 | 0 (0%) | 0 (0, 0) | NA (0, Inf) | <0.0001 | 5 (0.38%) | 1.04 (0.43, 2.51) | 1.2 (0.3, 4.8) | 0.8 |
| 2015 | 838 | 0 (0%) | 0 (0, 0) | NA (0, Inf) | <0.0001 | 0 (0%) | 0 (0, 0) | NA (0, Inf) | <0.0001 | 6 (0.72%) | 0.85 (0.38, 1.88) | 0.98 (0.2, 4.82) | 0.98 |
| 2017 | 514 | 0 (0%) | 0 (0, 0) | NA (0, Inf) | <0.0001 | 0 (0%) | 0 (0, 0) | NA (0, Inf) | <0.0001 | 4 (0.78%) | 0.85 (0.32, 2.27) | 0.99 (0.18, 5.34) | 0.99 |
| 2019 | 294 | 0 (0%) | 0 (0, 0) | NA (0, Inf) | <0.0001 | 0 (0%) | 0 (0, 0) | NA (0, Inf) | <0.0001 | 3 (1.02%) | 1.27 (0.41, 3.91) | 1.46 (0.25, 8.69) | 0.67 |
| 2022 | 170 | 5 (2.94%) | 4.13 (1.71, 10.02) | NA (0, Inf) | <0.0001 | 3 (1.76%) | 2.72 (0.87, 8.53) | NA (0, Inf) | <0.0001 | 3 (1.76%) | 2.37 (0.77, 7.32) | 2.74 (0.46, 16.26) | 0.27 |

| | | rtM41L | | | |
|---|---|---|---|---|---|
| **Survey round** | ***n*** | **Obs (%)** | **Prev. (95% CI)** | **Prev. ratio (95% CI)** | ***p*-value** |
| 2012 | 1223 | 4 (0.33%) | 1.52 (0.57, 4.05) | ref | ref |
| 2014 | 1308 | 5 (0.38%) | 1.01 (0.41, 2.48) | 0.67 (0.19, 2.29) | 0.52 |
| 2015 | 838 | 16 (1.91%) | 2.3 (1.42, 3.74) | 1.52 (0.51, 4.52) | 0.46 |
| 2017 | 514 | 10 (1.95%) | 2.16 (1.17, 3.99) | 1.42 (0.45, 4.52) | 0.55 |
| 2019 | 294 | 8 (2.72%) | 3.4 (1.72, 6.74) | 2.24 (0.68, 7.4) | 0.18 |
| 2022 | 170 | 3 (1.76%) | 2.48 (0.8, 7.7) | 1.63 (0.36, 7.3) | 0.52 |

Estimates were generated using Poisson regression with robust standard errors with survey round as a predictor variable. Multi-class refers to any combination of simultaneous resistance to any of the four drug classes. Generalized estimating equations with independent correlation structure to aid convergence for rare mutations were used. 95% confidence intervals indicate the Wald confidence interval around the mean value in each category. *p*-values that coefficients are different from 0 at the  level were calculated using the Wald method.
