## Appendix 3 for "Patterns of HIV-1 viral load suppression and drug resistance during the dolutegravir transition: a population-based longitudinal study"

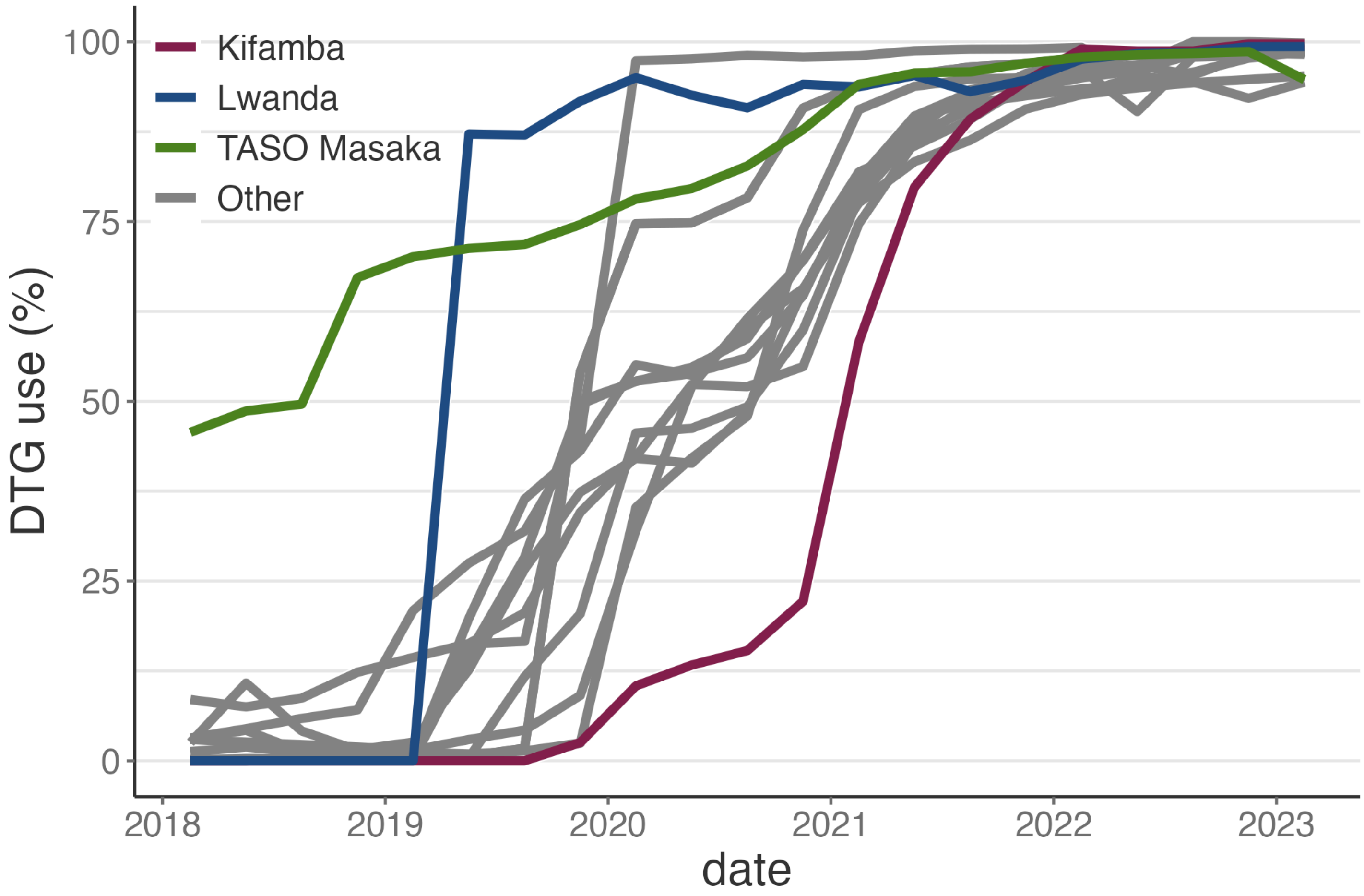


**Supplementary Figure 1: DTG-transition among clinics covering Rakai Community Cohort Study (RCCS) participants 2018-2022.** B) Proportion of PLHIV on DTG-based regimens at the top 15 clinics serving RCCS participants stratified by clinic.

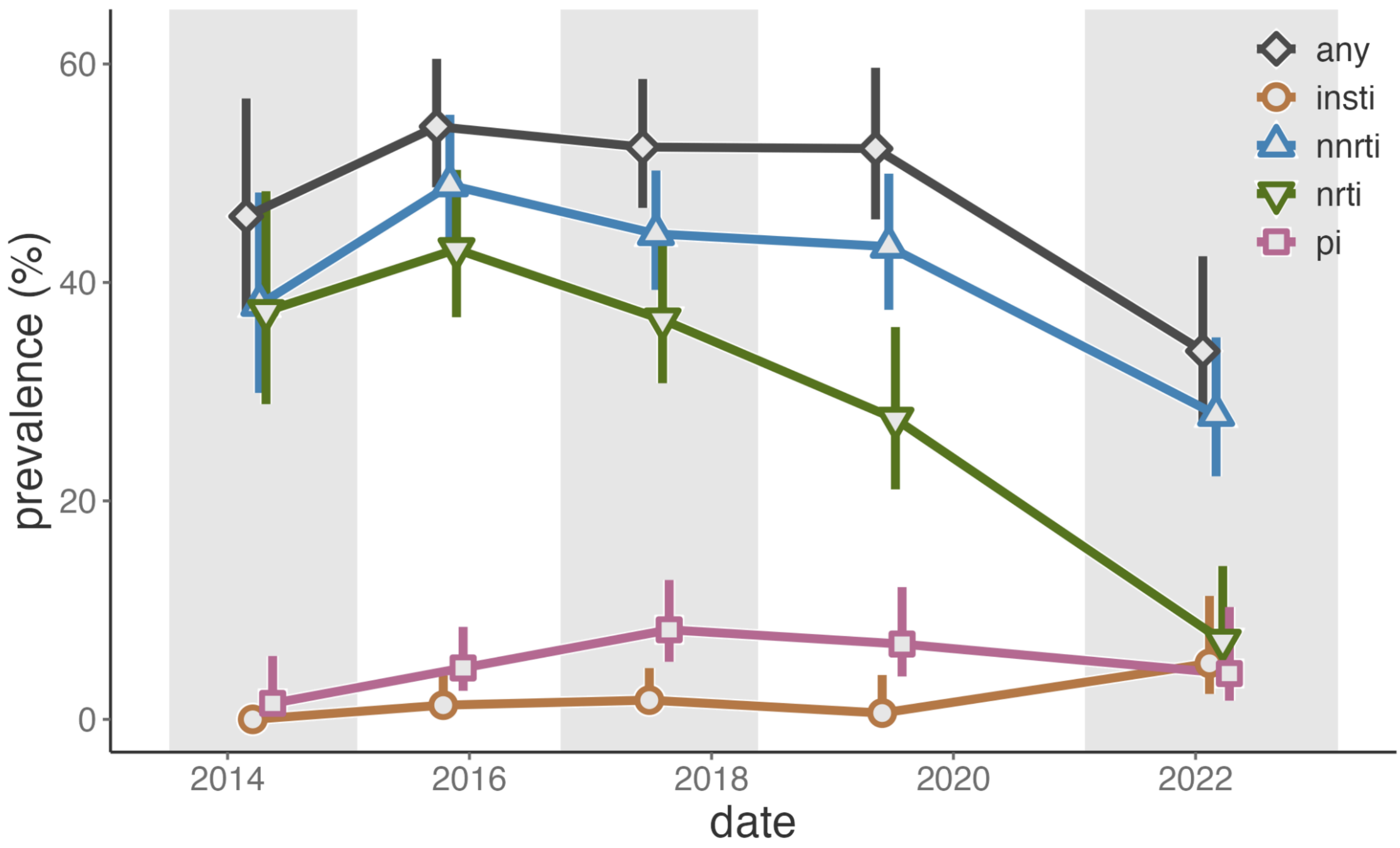


**Supplementary Figure 3: ART resistance among treatment-experienced viremic PLHIV in the Rakai Community Cohort Study using a 5%, 3 read variant calling threshold.** Prevalence of resistance to any drug class (grey), integrase strand transfer inhibitors (INSTIs, orange), non-nucleoside reverse transcriptase inhibitors (NNRTIs, blue), nucleoside reverse transcriptase inhibitors (NRTIs, green), and protease inhibitors (PIs, pink) by survey round among treatment-experienced viremic PLHIV. Vertical bars extend to the Wald 95% confidence intervals. Generalized estimating equations with correlation structure selection by Quasi Information Criterion value (Any: exchangeable, NNRTI: AR1, NRTI: independence, PI: exchangeable) or independence for outcome measures with less than 20 events (INSTI) were used to account for repeat participants across study rounds. Shading corresponds to the range of interview dates for alternating RCCS survey rounds.

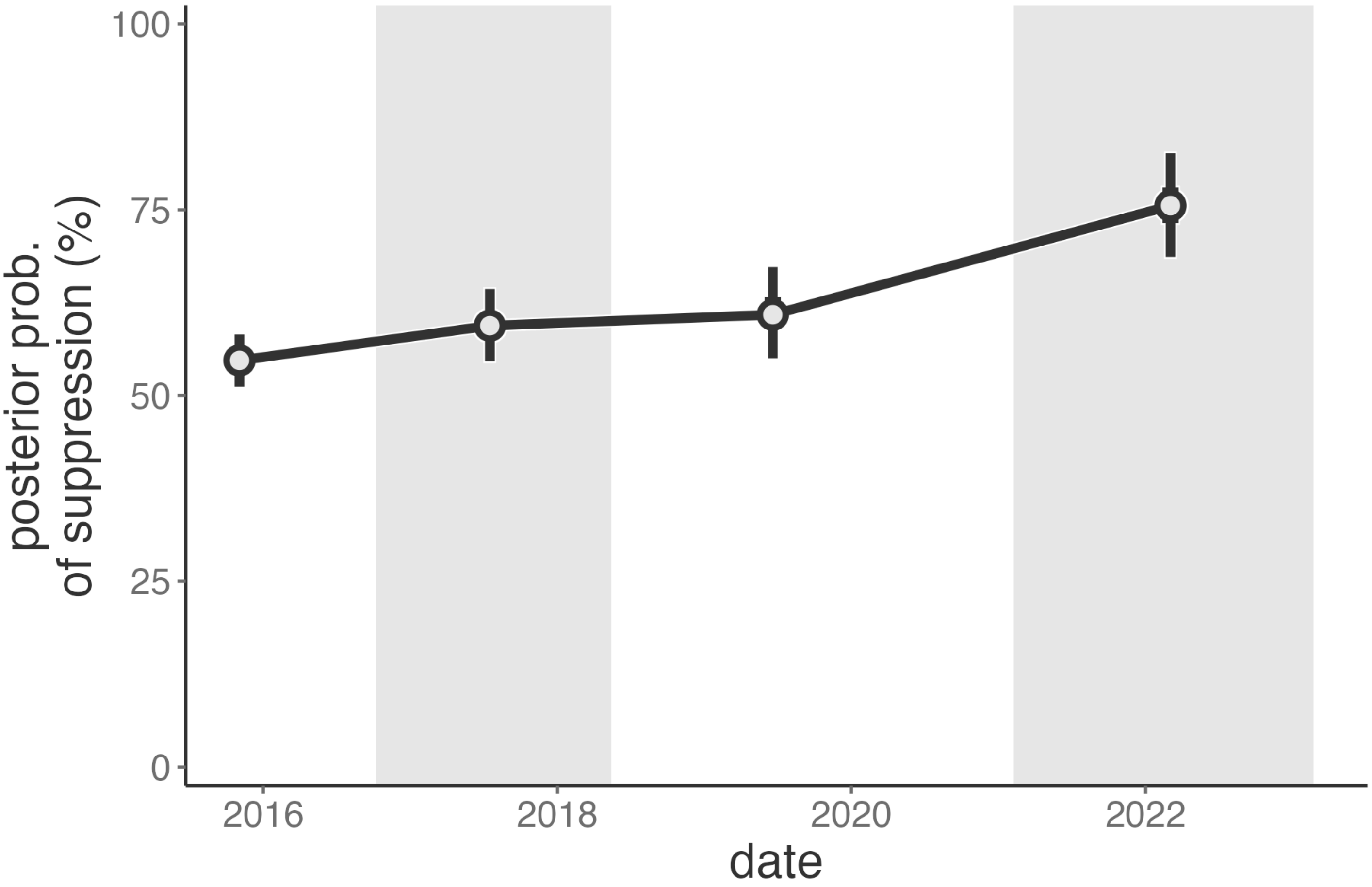


**Supplementary Figure 2: Probability of viral load suppression among Rakai Community Cohort Study participants who were viremic in the proceeding survey around.** Median value of the posterior distribution plotted as the central estimate and bars extend to the 50% and 95% highest posterior density. Estimates are plotted at the median date of the follow-up survey. Shading corresponds to the range of interview dates for alternating RCCS survey rounds.

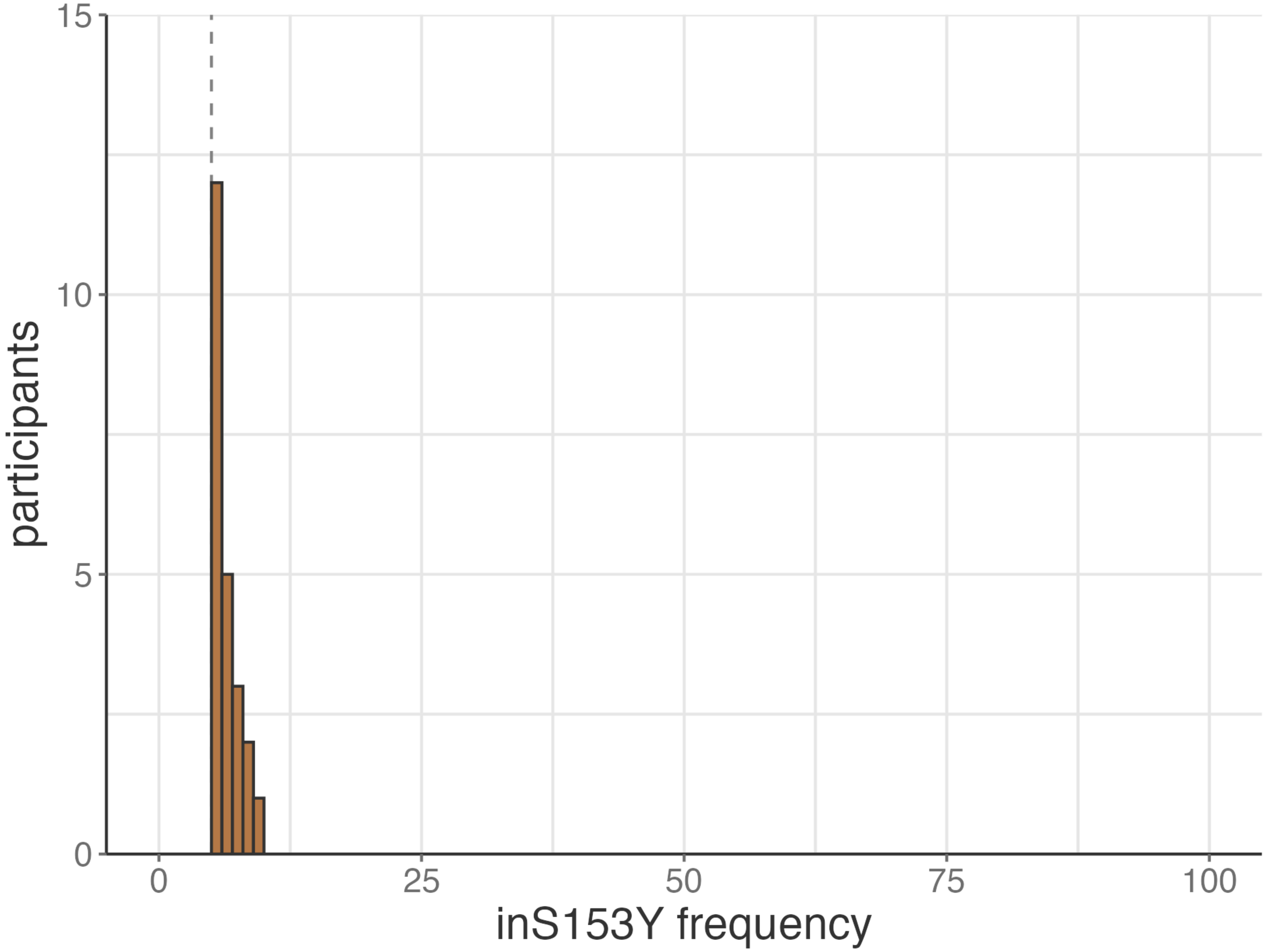


**Supplementary Figure 3: Within-host frequency of inS153Y among Rakai residents, 2022.** Vertical dotted line represents the 5% variant calling threshold.

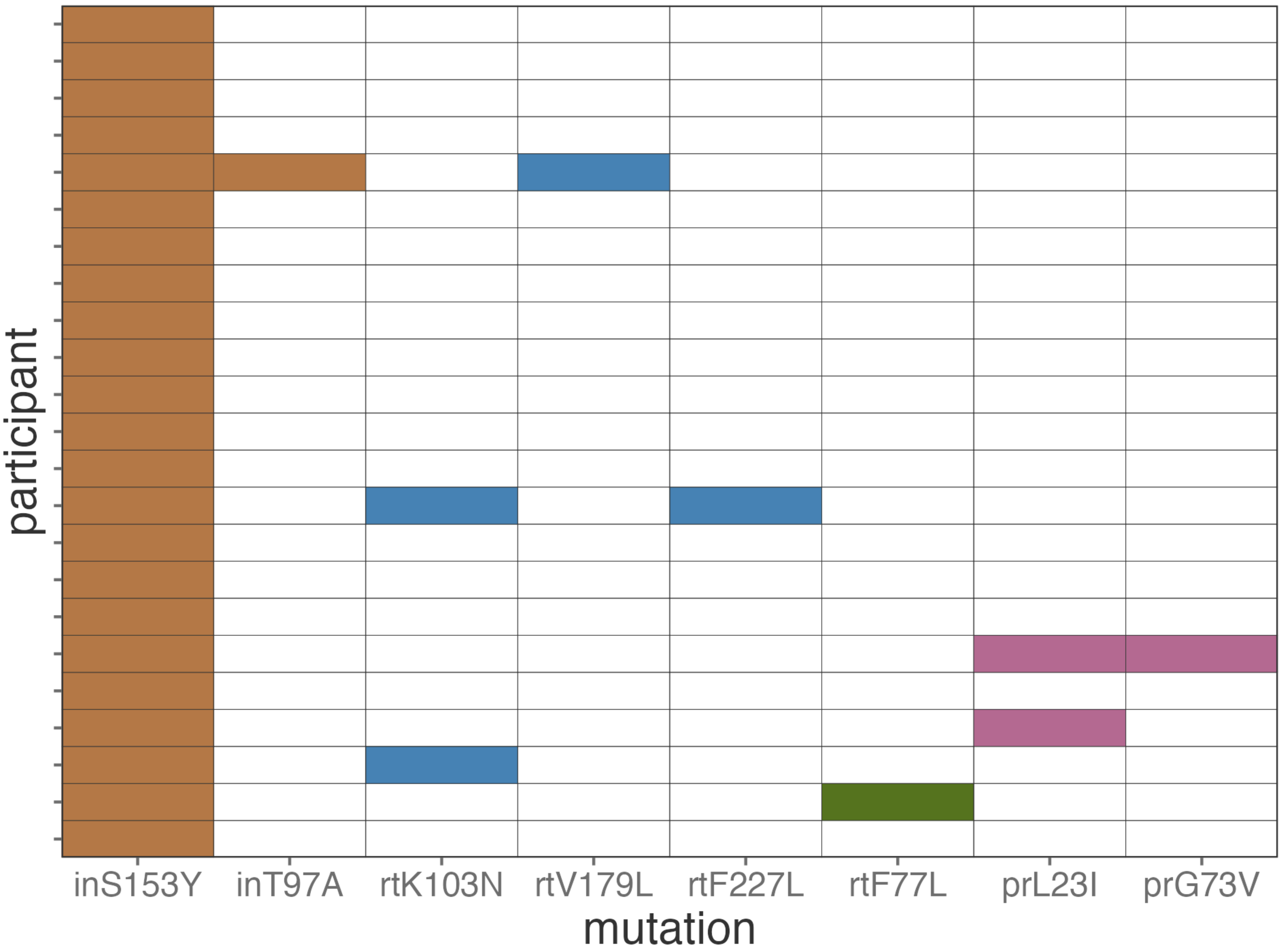


**Supplementary Figure 4: Co-occurrence of inS153Y with other major resistance mutations among Rakai residents, 2022.** Each row represents a distinct study participant. Filled in circles, colored by the class of drug to which a mutation confers resistance to (NNRTI: blue, NRTI: green, PI: pink), indicates mutation presence after application of our 5%/10 read threshold.
