## Appendix 1 for "Patterns of HIV-1 viral load suppression and drug resistance during the dolutegravir transition: a population-based longitudinal study"

### HIV Deep-sequencing and bioinformatic processing

HIV deep-sequencing from venous blood samples was performed through the Phylogenetics and Networks for Generalized HIV Epidemics in Africa consortium (PANGEA-HIV) using one of two methods as previously described[1–3]. Viral RNA extraction was performed using the QIAsymphony DSP Virus/Pathogen Kit. For amplicon-based deep-sequencing cDNA was generated using universal primers that span four overlapping regions of the HIV-1 genome in one-step reverse transcription PCR[4]. Sequencing using the Illumina MiSeq and HiSeq was conducted at the Wellcome Trust Sanger Institute. Alternatively, library preparation for the bait-capture protocol (veSEQ-HIV[5]) was performed with the SMARTer Stranded Total RNA-Seq v2-PicoInputMammalian and cDNA generated using in-house indexed primers. Libraries were pooled, cleaned with Agencourt AMPure XML, and hybridized to biotinylated 120-mer HIV-specific oligonucleotides (xGen Lockdown Probes, Integrated DNA Technologies). Following isolation with streptavidin-conjugated beads, libraries were PCR amplified and 350-600 base pair (bp) reads generated with the Illumina NovaSeq 6000 at the Oxford Genomics Centre.

Sequencing reads of viral and unknown origin were isolated using Kraken v.0.10.5-beta[6] with a database of human, bacterial, archaeal, viral and fungal genomes. After trimming of adaptors and low-quality bases with trimmomatic[7], reads were assembled into contigs using SPAdes[8] and metaSPAdes v.3.10[9]. Shiver v.1.5.7[10] was used to identify the most closely related reference sequence based on assembled contigs, generate complete sample-specific reference genomes, and align reads to sample-specific references. Samples were subtyped based on the subtype of the most closely related reference sequence.

To calculate pairwise genetic distances, a multiple sequence alignment of consensus sequences and HXB2 (GenBank: K03455.1) was generated using MAFFT v.7.490[11]. The polymerase region was extracted and any sequence with non-ambiguous (e.g. A, C, T, and G) coverage <95% was dropped using Numpy v.2.0.1[12] in Python v.3.12.4[13]. Codons with known drug resistance mutations were removed and Kimura 2-parameter[14] pairwise distances calculated in Python. For each subtype A1 and subtype D HIV sequence from PLHIV with the inS153Y mutation we identified the 10 most closely related Rakai sequences based on genetic distance. Phylogenetic trees of these A1 and D sequence sets were inferred using IQ-Tree v.2.2.2.7.[15]

### Imputation of 2012 survey round viral load measurements

Viral load measurements were not routinely collected for all participants with HIV in the 2012 survey round and we therefore randomly imputed viremia status among a subset of PLHIV in the 2012 survey round. As viral load measurements were nearly universally unavailable among treatment-experienced PLHIV in the 2012 survey round, we restricted any viral-load dependent outcome measures among populations including treatment-experienced PLHIV to the 2014 survey round and later. Because sequencing success depends on viral load we therefore impute viremia status (True/False) among by first calculating the probability of being viremic ($v_i = 1$) among those pre-treatment ($t_i = 0$) PLHIV ($h_i = 1$) in the 2012 survey round ($r_i = 2012$) on which viral load data were available ($q_i = 1$) stratified by whether deep-sequencing was successful for at least one drug class ($p_i = 1$). Specifically (removing the shared conditionals $t_i = 0,\ h_i = 1,\ r_i = 2012$ for simplicity):

$$P(v_i = 1 \mid p_i = 1,\ q_i = 1) = \frac{\sum_i^{n_{p=1,q=1}} v_i}{n_{p=1,q=1}},$$

$$P(v_i = 1 \mid p_i = 0,\ q_i = 1) = \frac{\sum_i^{n_{p=0,q=1}} v_i}{n_{p=0,q=1}},$$

where $n_{p=1,q=1}$ and $n_{p=0,q=1}$ are the number of pre-treatment PLHIV in the 2012 survey round with available viral load measurement on which deep-sequencing based resistance prediction was successful or unsuccessful for at least one drug class, respectively. Using these quantities we impute missing viremia status among those lacking viral load data as:

$$(v_i \mid p_i = 1,\ q_i = 0) \sim \text{Bernoulli}(P(v_i = 1 \mid p_i = 1,\ q_i = 1)$$

$$(v_i \mid p_i = 0,\ q_i = 0) \sim \text{Bernoulli}(P(v_i = 1 \mid p_i = 0,\ q_i = 1).$$

Among 2,641 self-reported pre-treatment PLHIV in the 2012 survey round viral load measurements are available for 1,606 (60.81%). Deep-sequencing was attempted on 2,586 (97.92%) and successful for at least one drug in 1825 (72.89%). A total of 792 (76.52%) participant-visits with missing viral load were imputed to be viremic and 243 (23.48%) imputed to be non-viremic and therefore not considered in our analyses of pre-treatment PLHIV.

**Longitudinal Bayesian Binomial logistic regression of paired viral load measurements**

We quantified the individual-level probability of viral load suppression in the subsequent RCCS survey ($s_{i,j}$) using data from pairs of participant-visits $j$ from the same individual $i$ in consecutive survey rounds. As we were here primarily interested in estimating the probability of achieving, as opposed to maintaining, suppression, we subset to viremic participant-visits ($v_{i,j} = 0$). Our model was therefore of the form

$$(s_{i,j} \mid v_{i,j} = 0) \sim \text{Binomial}(\mu_{i,j})$$
$$\text{logit}\left(\mu_{i,j}\right) = \alpha_0 + \alpha_i + \sum_{k=1}^{n_c} X_{i,j}^k \beta_k .$$

Where $\alpha_i$ represents individual-level effects to account for the fact that participants may contribute to multiple surveys (although contribute at most once to each individual survey), $X_{i,j}^k$ are $1 \times z_k$ dimensional row vectors for each of $n_c$ epidemiological covariates and $\beta_k$ are $z_k \times 1$ column vectors of fixed effect coefficients. Specifically, we model categorical covariates for survey round (2014/2015/2017/2019), age category ((14,24]/(24,34]/(34,49]), community type (inland/fishing), and sex (male/female). For all models we use diffuse priors and include all levels of categorical covariates in $X_{i,j}^k$ while enforcing a sum-to-zero joint multivariate normal prior on $\beta_k$ such that all marginal distributions are $N(0, 1)$ to main identifiability and equivalent prior distributions for all levels. Specifically,

$$\alpha_0 \sim N(0, 1)$$
$$\alpha_i \sim N(0, \sigma)$$
$$\sigma \sim \text{Half-Cauchy}(0, 1)$$
$$\beta_k \sim \text{stz-MVN}(0, 1).$$

To quantify the impact of resistance to NNRTIs and NRTIs on viral load suppression in the subsequent survey round we subset the analysis to the participant-visits with available sequence data, $p_{i,j} = 1$. We allow the effect of resistance on suppression to vary by survey round, specifically,

$$\left(s_{i,j}^a \mid p_{i,j} = 1, s_{i,j} = 0\right) \sim \text{Binomial}\left(\mu_{i,j}^a\right)$$
$$\text{logit}\left(\mu_{i,j}^a\right) = \alpha_0 + \alpha_i + \sum_{k=1}^{n_c} X_{i,j}^k \beta_k + \sum_{r=1}^{n_r} R_{i,r}^a \delta_r^a$$
$$\delta_r^a \sim \text{stz-MVN}(0, 1),$$

where $R_{i,r}^a$ are $1 \times 2$ dimensional row vectors representing the interaction between resistance to drug class $a$ for participant $i$ in survey round $r$. As above, $\delta_r^a$ are $2 \times 1$ dimensional column vectors with fixed effects coefficients for the absence or presence of resistance to drug class $a$ in round $r$.

Further, to aid in the calculation of our target quantities (Generated quantities) we modelled the probability of resistance to NNRTIs and NRTIs ($y^a_{i,j}$) among those with available sequence data, regardless of participation in the subsequent survey, as a function of survey round, age, sex, and community type.

$$\left(y^a_{i,j} | p_{i,j} = 1\right) \sim \text{Binomial}\left(\pi^a_{i,j}\right)$$

$$\text{logit}\left(\pi^a_{i,j}\right) = \alpha^\pi_0 + \alpha^\pi_i + \sum_{k=1}^{n_c} X^k_{i,j} \beta^\pi_k$$

$$\alpha^\pi_0 \sim N(0, 1)$$

$$\alpha^\pi_i \sim N(0, \sigma^\pi)$$

$$\sigma^\pi \sim \text{Half-Cauchy}(0, 1)$$

$$\beta^\pi_k \sim \text{stz-MVN}(0, 1).$$

**Generated quantities**

Based on our logistic regression models defined above, we first quantified the probability of viral load suppression in the subsequent round given viremia in the 2012, 2015, 2017, and 2019 surveys. To account for the fact that not all viremic participants in a given survey participate in the following survey, we make the simplifying assumption that follow-up is independent of suppression within strata of survey round, age, sex, and community type. We then estimate $\mu_g$ within epidemiological sub-groups $g$ defined by survey round, age, sex, and community type based on the design matrix used in the model above and use post-stratification to weight the strata-specific estimates by the total number of viremic participants in each strata, regardless of participation in the following round, $w_g$, and sum over the strata in each round to generate round-specific estimates $\tilde{\mu}_r$. Specifically,

$$\mu_g = \text{inverse-logit}\left(\alpha_0 + \sum_{j=1}^{n_c} X^j_g \beta_j\right)$$

$$\tilde{\mu}_r = \frac{\sum_{g \in r} w_g \mu_g}{\sum_{g \in r} w_g}$$

Next, we quantified the probability of viral load suppression in the subsequent round given viremia in the 2012, 2015, 2017, and 2019 surveys stratified by NNRTI and NRTI resistance. We first calculate the estimated probability of suppression within sub-groups, conditional on the presence and absence of resistance,

$$\mu^{y_a=0}_g = \text{inverse-logit}\left(\alpha_0 + \sum_{j=1}^{n_c} X^j_g \beta_j + \beta^\alpha_{r_g}[1]\right)$$

$$\mu^{y_a=1}_g = \text{inverse-logit}\left(\alpha_0 + \sum_{j=1}^{n_c} X^j_g \beta_j + \beta^\alpha_{r_g}[2]\right),$$

where $\beta^\alpha_{r_g}[1]$ and $\beta^\alpha_{r_g}[2]$ represent the first and second elements of the $\beta^\alpha$ vectors corresponding to the survey rounds for sub-group $g$, respectively. Next, we calculate the estimated probability of resistance within each sub-group, conditional on the availability of sequence data,

$$\pi^a_g = \text{inverse-logit}\left(\alpha^\pi_0 + \alpha^\pi_i + \sum_{k=1}^{n_c} X^k_g \beta^\pi_k\right).$$

Finally, by making the additional simplifying assumption that within-sub-groups defined by survey round, age, sex, and community type the probability of resistance is the same between those viremic PLHIV with and without available sequence data,

$P\left(y^{a}_{i,j} = 1 \mid p_{i,j} = 1,\ v_{i,j} = 1\right) = P\left(y^{a}_{i,j} = 1 \mid p_{i,j} = 0,\ v_{i,j} = 1\right)$, we can estimate the total number of viremic participants with resistance in each sub-group. Using this estimate, we arrive at the estimated probability of suppression in each survey round, conditional on resistance status,

$$\hat{\mu}_{r}^{y_a=0} = \frac{\sum_{g\in G_r}\left(1-\pi^{a}_{g}\right)w_g\mu_g^{y_a=0}}{\sum_{g\in G_r}\left(1-\pi^{a}_{g}\right)w_g}$$

$$\hat{\mu}_{r}^{y_a=1} = \frac{\sum_{g\in G_r}\left(\pi^{a}_{g}\right)w_g\mu_g^{y_a=1}}{\sum_{g\in G_r}\left(\pi^{a}_{g}\right)w_g},$$

where $G_r$ is the set of all epidemiological sub-groups associated with round $r$. Parameter inference was conducted in CmdStanR v.0.8.1[16] with 2,500 iterations of warm-up and sampling for four chains and posterior distributions were summarized using HDInterval v.0.2.4[17].

14 Kimura M. A simple method for estimating evolutionary rates of base substitutions through comparative studies of nucleotide

sequences. *J Mol Evol* 1980; **16**: 111–20.

15 Minh BQ, Schmidt HA, Chernomor O, *et al.* IQ-TREE 2: New Models and Efficient Methods for Phylogenetic Inference in the Genomic Era. *Mol Biol Evol* 2020; **37**: 1530–4.

16 Gabry J, Češnovar R, Johnson A. cmdstanr: R Interface to ‘CmdStan’. 2024. https://mc-stan.org/cmdstanr/.

17 Meredith M, Kruschke J. HDInterval: Highest (Posterior) Density Intervals}. 2022. https://CRAN.R-project.org/package=HDInterval.
